# Assessing Respiratory, Cardiac and Neural Interactions During Rest and Breath-Focus in Novice and Expert Meditation Practitioners

**DOI:** 10.1101/2025.06.29.25330504

**Authors:** Javier R. Soriano, Angeliki-Ilektra Karaiskou, Julio Rodriguez-Larios, Carolina Varon, Nazareth Castellanos, Kaat Alaerts

## Abstract

**Background:** Breath-focused meditation is recognized for its potential to enhance autonomic regulation and attentional control. However, most research focuses on the study of isolated physiological domains, limiting the understanding of how neural and cardiorespiratory systems interact dynamically to support self-regulatory processes.

**Methods:** EEG, ECG, and respiratory activity were recorded in 54 participants (27 expert meditators, 27 novices) during periods of rest and breath-focused meditation. Respiratory rate, heart rate, and heart rate variability changes were assessed, as well as directional information flow computed between alpha-band neural oscillations, cardiac, and respiratory signals using transfer entropy. Additionally, cross-frequency dynamics were studied as proposed by a recent theory of brain-body coupling.

**Results:** Both groups exhibited reduced respiratory rates during meditation, with experts showing the lowest rates. Parasympathetic tone, assessed while controlling for respiratory confounds, was higher in experts across conditions. Transfer entropy analyses revealed distinct patterns of neurovisceral integration: novices demonstrated stronger bottom-up cardiac and respiratory influences on posterior cortical alpha activity, whereas experts showed enhanced top-down alpha to respiration coupling, particularly at frontocentral midline and right prefrontal sites. Cross-frequency ratio analyses revealed a reduction of the 8:1 harmonic alpha: heart rate ratio during meditation compared to rest in experts.

**Conclusions:** This study advances the understanding of how meditation expertise shapes dynamic brain-body interactions, emphasizing the experience resulting shift from reactive to proactive interoceptive control aligned with predictive processing theories. By integrating measures across physiological systems and their bidirectional interplay, these findings further enrich theoretical accounts of embodied cognition and autonomic regulation, and inform the development of novel tools to understand and facilitate meditation practices, such as combined neuro-and biofeedback protocols.

## Introduction

The increasing availability of mindfulness-related media, resources, and apps has led to a surge in the number of practitioners (Davies et al., 2024). While various meditation traditions exist (Brandmeyer et al., 2019; Lutz et al., 2015; Sparby & Sacchet, 2022), the secularization of mindfulness within Mindfulness-Based Stress Reduction (MBSR) programs (Kabat-Zinn, 2013) has popularized focused attention meditation as one of the most widely practiced and studied techniques (Baminiwatta & Solangaarachchi, 2021; Galante et al., 2023; Goldberg et al., 2022). Focused attention meditation involves sustaining attention on a chosen object, such as bodily sensations, and redirecting focus when distractions arise (Kabat-Zinn, 2013). This practice benefits numerous cognitive functions, including attention regulation (Ainsworth et al., 2013; Moore et al., 2012; Yoshida et al., 2020), metacognition (Baird et al., 2014), stress management (Chiesa & Serretti, 2009; Goldberg, 2022; Kriakous et al., 2021; Rosenkranz et al., 2016) and emotion regulation (Doll et al., 2016). Ultimately, focused attention meditation aims to cultivate equanimity, promoting present moment awareness (Chems et al., 2025). Its psychological and mental health advantages have been extensively studied (Chan et al., 2017; Galante et al., 2021; Gallant, 2016; Prätzlich et al., 2016; Quach et al., 2016) and, given their therapeutic potential, research continues to explore focused attention and mindfulness-based training tools for both general and clinical populations.

Previous research on the neural correlates of focused attention meditation has predominantly examined alpha oscillations, despite effects documented across the broader frequency spectrum (Lieberman et al., 2024). Mindful attention towards body sensations (e.g., breath related) is proposed to modulate alpha activity through enhanced top-down control, via engagement of prefrontal areas, which regulate thalamocortical circuits, exerting attentional control and filtering of irrelevant sensory information (Kerr et al., 2013). Alpha oscillations are particularly relevant due to their prominence and stability, especially at rest with closed eyes (Mazaheri et al., 2014). Additionally, they are tightly associated with attention (Xia et al., 2024), arousal (Kawashima et al., 2024; Mierau et al., 2017), task-unrelated inhibition (Chapeton et al., 2019; Haegens et al., 2011; Jensen & Mazaheri, 2010) and mind wandering (Brandmeyer & Delorme, 2018; Kam et al., 2022; Rodriguez-Larios et al., 2021). Although many studies report increased alpha power during focused attention meditation relative to rest and control tasks (Lee et al., 2018; Lieberman et al., 2024; Lomas et al., 2015), others indicate decreases, particularly in experienced meditators (Rodriguez-Larios et al., 2021; Rodriguez-Larios, Faber, et al., 2020; Saggar et al., 2012). Notably, reductions in alpha peak frequency after Mindfulness-Based Stress Reduction (MBSR) interventions also suggest decreased arousal levels (Rodriguez-Larios et al., 2024). In a study involving participants analyzed in the current work, breath-focused meditation reduced alpha power and peak frequency relative to rest only in experts, while novices showed increased frontal alpha during mind-wandering episodes (Rodriguez-Larios et al., 2021). Additionally, focused attention meditation typically increases neural complexity (Kakumanu et al., 2018; Lu & Rodriguez-larios, 2022; Walter & Hinterberger, 2022), whereas expert meditators exhibit decreased neural complexity at baseline, suggesting a trait effect (Atad et al., 2023).

There is increasing recognition that meditation induces changes not only at the neural, but also visceral levels, including respiratory and cardiac activities. In addition to its central role in breath-focused practices, respiration has gained significant attention in meditation research due to its role in interoception and stress regulation (Harrison et al., 2021; Kabat-Zinn, 2013). Furthermore, respiration patterns influence several emotional (Ashhad et al., 2022), neural, and cognitive processes (Allen et al., 2022; Azzalini et al., 2019; Belli & Fischer, 2024; Braendholt et al., 2023; Goheen et al., 2023, 2024; Herrero et al., 2018; Jelinčić et al., 2022; Kluger et al., 2021, 2024; Lewis-Healey et al., 2024; Li et al., 2024; Maric et al., 2020; Nakamura et al., 2024; Signorelli et al., 2022; Sterling, 2018; Zelano et al., 2016). Although breath-focused meditation practices do not typically instruct specific breathing patterns (Bentley et al., 2023), reductions in respiration rate (RR) during mindfulness meditation compared to rest and control conditions (Ahani et al., 2014; Atchley et al., 2016) and at rest after mindfulness practices are commonly observed (Dietrich & Bidart, 2024; Karunarathne et al., 2024; Rusinova et al., 2024; Wielgosz et al., 2016). Crucially, respiratory activity strongly influences cardiac function; a reduced RR can enhance vagal activity, decreasing sympathetic arousal and facilitating emotion regulation (Balzarotti et al., 2017; Prescott & Liberles, 2022), especially in expert meditators with lower RR further becoming a trait (Gerritsen & Band, 2018; Wielgosz et al., 2016).

Consistent with the systemic view that physiological functions interact dynamically (Bartsch et al., 2015), the relationship between cardiac and respiratory activities with brain function has been explored across various contexts, (Adelhöfer et al., 2020; Candia-Rivera et al., 2024; Catrambone et al., 2024; Corcoran et al., 2023; de Zambotti et al., 2018; Hsueh et al., 2023; Lovelace et al., 2024; Sargent et al., 2024; Silvani et al., 2016). Although growing evidence supports the notion of functional neurovisceral interdependence during meditation, such as the alteration of bidirectional brain-heart interplay under the elicitation of mental stress (Candia-Rivera et al., 2023; Pernice et al., 2021; Zanetti et al., 2019), most contemplative research has examined physiological systems independently (Bailey et al., 2024; Lumma et al., 2015), thus providing limited insight into interactions between physiological signals, or restricting analyses to correlation-based metrics (Anurag et al., 2023; Gao et al., 2023; Kim et al., 2013; Krishna & Prasanna, 2023; Sik et al., 2017; Tang et al., 2009; Wong et al., 2022). A smaller number of studies has explored neurovisceral interactions during meditation (Jiang et al., 2020; Soriano et al., 2024; Wang et al., 2024). For instance, Soriano et al., (2024) investigated neural and cardiac cross-frequency dynamics during breath-focus and rest while other studies investigated heartbeat-evoked potentials (HEP) (Jiang et al., 2020; Wang et al., 2024). Jiang et al. (2020) reported HEP amplitude differences in experienced meditators during meditation compared to rest, and at rest compared to controls, particularly over frontocentral regions, suggesting heightened interoceptive awareness. In contrast, Wang et al. (2024) did not find amplitude differences between meditators and controls at rest, but observed a more posterior HEP distribution in experts, pointing to altered cardiac signal processing. So far, the mentioned studies either look only at novices (Soriano et al., 2024) or only at experts during breath focus or a mix of techniques (Jiang et al., 2020), or both groups but only at rest (Jiang et al., 2020; Wang et al., 2024), thus not accounting for experience-related differences during meditation practice.

Regarding neurorespiratory interactions, a study by Herrero et al. (2018) found increased coherence between neural (intracranial EEG) and respiratory activity during a breath counting task in trials with a correct counting performance (classified as attentive trials), although alpha power did not differ between correct and incorrect (non-attentive) trials. While pure attention to the breath without a counting component remains uninvestigated in terms of alpha–respiratory signal interactions, several studies have shown that breathing manipulations (particularly slow, deep, or nasal breathing) can influence neural activity, with the alpha rhythm emerging as a potential marker of the regulatory and affective effects of such practices (see Jelinčić et al., 2022). For instance, Zelano et al. (2016) found that nasal breathing entrains limbic neural activity (specifically in the theta band), modulating memory recall and suggesting that respiration rhythmically gates limbic processing. Recent theoretical frameworks further support the role of slow breathing in shaping neural dynamics. Braendholt et al. (2023) propose that neural-respiratory coupling reflects a hierarchical organization of brain rhythms that synchronize with the timing of slow nasal breathing, whereby temporal alignment between internal bodily rhythms and neural activity are optimized. This entrainment is thought to facilitate sensorimotor integration and interoceptive inference, both relevant processes in contemplative and breath-focused practices.

With respect to potential mechanisms underlying oscillatory interactions between different physiological systems, the Binary Hierarchy Brain Body Oscillation Theory (Klimesch, 2018) proposes that brain and body rhythms interact dynamically, forming cross-frequency harmonic and non-harmonic relationships that either facilitate or prevent cross-frequency (phase)coupling. This framework suggests that two oscillators can phase-couple effectively when their frequencies maintain a harmonic, integer-based ratio, for example: alpha (12 Hz) and theta (6 Hz) oscillations in a 2:1 relationship (Klimesch, 2018). In contrast, non-harmonic relationships, such as those based on the irrational phi ratio (1.618:1), create highly desynchronized states, potentially precluding spurious coupling (Pletzer et al., 2010). Research shows that neural cross-frequency dynamics are modulated by cognitive demands, with enhanced harmonic phase coupling during effortful tasks (Palva et al., 2005; Rodriguez-Larios & Alaerts, 2019; Sauseng et al., 2008; Siebenhühner et al., 2016). Conversely, mindfulness meditation is associated with reduced alpha–theta harmonic coupling and increased non-harmonic interactions, both in novices and experienced meditators (Rodriguez-Larios, Faber, et al., 2020; Rodriguez-Larios & Alaerts, 2019). These shifts also appear early during MBSR training (Rodriguez-Larios, Wong, et al., 2020), while mind-wandering episodes during meditation show stronger harmonic alpha–theta coupling, suggesting greater involvement of executive and memory processes than breath-focus self-reported episodes (Rodriguez-larios & Alaerts, 2021). Importantly, the theory extends beyond brain rhythms to include visceral oscillators such as cardiac activity. A typical harmonic neuro-cardiac ratio is proposed at 8:1, for instance, with 10 Hz alpha and 1.25 Hz heart rate (Klimesch, 2018). While neural-visceral cross-frequency dynamics have recently gained interest (Kluger & Gross, 2021; Rassi et al., 2019; Richter et al., 2017; Sargent et al., 2024; Soriano et al., 2024), they remain underexplored in meditation research. Recently, Soriano et al. (2024) demonstrated that heart rate modulations during breath-focused meditation and an arithmetic task produce distinct alpha: heart rate cross-frequency ratio profiles, highlighting task-dependent shifts in neural-cardiac cross-frequency relationships. Specifically, during the arithmetic task, a higher incidence of a cluster of ratios encompassing the 8:1 harmonic cross-frequency relationship was present, an alignment proposed to enable coupling among neural and cardiac rhythms under heightened cognitive effort.

In the current study, a mixed analytical approach is combined for the study of respiratory, neural, and cardiac oscillatory dynamics during rest and breath focus attention meditation in both novice and expert practitioners. On the one hand, individual physiological system differences across the two groups and conditions are assessed and analyses extended to assess bidirectional influences between the oscillatory profiles with information dynamics techniques. On the other hand, the cross-frequency interactions framework is employed in order to elucidate putative mechanisms that could explain these oscillatory network interactions. By integrating cross-frequency coupling analyses with measures of information flow, this study offers a comprehensive perspective, from both a theory-driven framework on oscillatory coordination and a data-driven approach serving as complementary investigations of the same phenomena.

## Methods

### Participants

From the initial 63 participants recruited for the study, 5 participants were excluded due to issues during data acquisition and 4 additional participants were excluded due to poor data quality. Amongst the remaining 54 participants included in the present work, 27 had no prior meditation experience (14 females, mean age 46.78 years, age range 27-64) and 27 were expert meditators (16 females, mean age 47.33 years, age range 29-67) with at least three years of expertise in either Mindfulness, Zen or Vipassana meditation. Informed consent forms and study design were approved by the Social and Societal Ethics Committee (SMEC) of KU Leuven, in accordance with the Declaration of Helsinki (dossier no. G-2019 09 1747). Participants were compensated for their participation with 8 € per hour in addition to travel costs.

### Design and Tasks

Participants took part in three conditions: a rest period, a breath-focused meditation practice, and a probe-caught mind-wandering focus task. This report focuses on the rest and meditation conditions, while the mind-wandering task is detailed elsewhere (Rodriguez-Larios et al., 2021). While at rest for 5 minutes, participants were required to keep their eyes closed and remain still, while not falling asleep and allowing spontaneous thoughts to arise. The meditation session lasted 10 minutes, of which only the last 5 minutes were selected for further analyses, and was preceded by a set of instructions aligning with MBSR practices (Kabat-Zinn, 2013): ‘Sit in a comfortable posture that embodies dignity, keeping the spine straight and letting your shoulders drop. Close your eyes and allow your attention to gently align to the sensation of breathing. You can focus on the part of your body where you feel your breath most clearly (for example: nostrils, belly, chest…). Every time you notice that your mind has wandered off your breath, notice what it was that carried you away, and then gently bring your attention back to the sensations associated with your breath’ (Kabat-Zinn, 2013; Rodriguez-Larios et al., 2021).

### Data acquisition and pre-processing

#### Recordings

Continuous electroencephalographic (EEG), electrocardiographic (ECG) and respiration recordings were simultaneously obtained during the rest and meditation conditions using the Nexus-32 system (Mind Media, The Netherlands) in combination with BioTrace software (version 2015a). EEG was continuously recorded with a cap fitted with 22 electrodes, including two references and one ground, positioned according to the international 10–20 system. EEG signals were amplified with a unipolar amplifier and sampled at 512 Hz. ECG was continuously monitored using a bipolar two-lead setup, with electrodes positioned beneath the left ribcage and just below the right clavicle; the EEG cap’s ground electrode served as the reference. The ECG signal was amplified through a bipolar amplifier and sampled at 256 Hz. Respiration was simultaneously recorded with an elastic belt secured around the thoracic region right below the chest.

#### EEG pre-processing

EEG data were processed using custom MATLAB scripts together with EEGlab (Delorme & Makeig, 2004) and FieldTrip (Oostenveld et al., 2011) functions. In order to match the data length of both conditions, the last 5 minutes of the meditation practice were selected, similar to previous work (Rodriguez-Larios et al., 2021), and then both conditions were pre-processed equally. First, the initial and final two seconds of each recording were removed from all physiological modalities. The data, originally sampled at 512 Hz, were then downsampled to 256 Hz to match the ECG sampling rate. A high-pass filter at 1 Hz and a low-pass filter at 40 Hz were applied using the *pop_eegfiltnew* function to eliminate slow drifts and high-frequency noise, respectively. Line noise (50, 100, and 150 Hz) was then attenuated using a notch filter. Noisy or flat channels were automatically identified using a combination of flat-line detection and correlation-based rejection (threshold set at 0.5). Removed channels were then interpolated using spherical splines. The data were then re-referenced to the common average. Independent component analysis (ICA) was thereafter performed using the *runica* algorithm, with the number of components set to the count of non-interpolated electrodes minus one to maintain data rank integrity. Artifact components, including those related to ocular, muscular, cardiac, or channel noise, were automatically identified and rejected using the *iclabel* function (Pion-Tonachini et al., 2019) with a probability cutoff of 80%.

#### ECG pre-processing

ECG signals were processed using custom MATLAB scripts. The raw ECG data were filtered using a zero-phase, 4th-order Butterworth bandpass filter with a passband of 1-40 Hz. filter. R-peaks within the QRS complex were automatically detected and annotated with the MATLAB toolbox R-DECO (Moeyersons et al., 2019) and corrected by visual inspection.

#### Respiration pre-processing

The data were smoothed using the *movmean* function with a window size factor of 0.4. After smoothing, the data were then standardized using z-scoring to normalize amplitude. A zero-phase filter was applied to the standardized signal with stopbands set at 0.05 Hz and 1 Hz, and a passband of 0.06-0.9 Hz. The power spectral density (PSD) of the preprocessed respiration signal was estimated using Welch’s method (*pwelch*), with a 40-second window, 50% overlap, and 1024-point FFT resolution. Finally, the respiration signal was resampled to 256 Hz to match that of the EEG and ECG data.

### Feature extraction

#### Cardiorespiratory analyses

The peak respiration frequency was identified by locating the maximum power in the estimated PSD, and the corresponding frequency value was extracted as the dominant respiration rate for each participant and condition. Heart rate time series were derived from the detected R-peaks in the ECG signal. First, inter-beat intervals (IBIs) were calculated by taking the difference between consecutive R-peak indices. The cumulative sum of IBIs provided the corresponding time points for each beat. Instantaneous heart rate values were then computed by taking the inverse of each IBI (in Hz). To obtain a continuous heart rate time series, the instantaneous values were interpolated onto a regular time grid at a sampling frequency of 1 Hz. Lastly, the mean heart rate was calculated per participant and condition.

Regarding the estimation of heart rate variability (HRV), the high-frequency (HF) HRV component (0.15-0.4 Hz) aligns with the respiratory sinus arrhythmia (RSA) and serves as a parasympathetic marker under typical resting breathing rates (Shaffer & Ginsberg, 2017). However, in practices such as breath-focused meditation, where respiration often slows down, breathing frequencies can fall below the HF range, shifting power toward the low-frequency component (LF) and compromising the reliability of both frequency components as markers of ANS tone (Laborde et al., 2017; Quintana & Heathers, 2014; Ritz, 2024a, 2024b; Shaffer & Ginsberg, 2017). This might explain why some studies focusing on frequency analyses of HRV during meditation practices have found no reliable parasympathetic increases, potentially due to not adequately controlling for respiration rate when interpreting power in the HF (Brown et al., 2021). Considering that meditation can in fact increase HRV (Gerritsen & Band, 2018), alternative HRV markers are necessary for slow-breathing meditation studies as the present one. To estimate HRV indices independently of respiratory influences, the method described by Varon et al. (2019) was applied, which uses orthogonal subspace projection (OSP) to decompose HRV into respiratory and residual components, with the former, here on denominated Px, reflecting a respiration-adjusted measure of parasympathetic modulation.

#### Cross-frequency ratio analyses

Estimation of transient alpha peak frequency and heart rate closely followed Soriano et al (2024). The cleaned EEG data were segmented into 1-second epochs, and epochs with an absolute amplitude exceeding 50 µV were excluded. Then, alpha peak frequencies (8–14 Hz) were extracted from 1-second EEG epochs using short-time Fourier transforms and peak detection, resulting in time series of alpha peaks per condition, participant, and electrode. Cross-frequency ratios between alpha frequency and heart rate were computed by dividing the alpha time series by the previously calculated heart rate time series in a second by second basis. For each possible alpha: heart rate ratio, ranging between 4 and 24 in steps of 0.5, the percentage of epochs displaying it was calculated per condition, participant and electrode, thus providing a distribution of the cross-frequency ratio profile.

#### Information dynamics analyses

To investigate bidirectional and cross-modal interactions between neural, cardiac, and respiratory systems, four transfer entropy (TE) measures were computed: 1) from alpha-band neural activity to HRV, 2) from HRV to alpha activity, 3) from alpha activity to respiration, and 4) from respiration to alpha activity. All TE calculations were performed within a unified framework based on multivariate autoregressive (VAR) modeling, which captures the temporal dynamics between pairs of physiological signals. For each pairing, the driver (source) and target (receiver) time series were modeled jointly, with the optimal VAR model order selected using Akaike information criterion, setting a maximum order of 20. This approach enables the quantification of directed information flow, allowing the assessment of how activity in one system predicts future changes in another. To that end, in order to extract the alpha power time series from the EEG and to compute the information flow between the alpha band and respiration as well as heart rate, the procedure described in Catrambone et al. (2021) was followed. The power spectral density (PSD) for each EEG electrode was computed using the Welch method, employing a 2-second Hamming window with a 0.25-second sliding step. This approach generated a time-frequency representation sampled at 4 Hz. A bandpass filtering rule was applied to the resulting PSD time series, maintaining only frequency components in the 8–14 Hz alpha band range. Similarly, both HRV and respiration signals were resampled to 4Hz and then processed to extract their respective power envelopes, ensuring consistent representation across all modalities. For each transfer entropy (TE) pair; alpha to HRV, HRV to alpha, alpha to respiration, and respiration to alpha, time series were mean-centered. Each VAR model was then fitted to capture the temporal dependencies between each driver and target pair. TE was calculated as the reduction in uncertainty of the target signal (**y**) achieved by including past information from the driver signal (**x**), beyond the predictive contribution of **y**’s own history. Specifically, TE was computed from conditional entropies derived from the VAR model parameters, providing a robust, model-based measure of directed information flow. For the TE calculation, a lag length of 10 was used to capture past dependencies in the time series as described in Morales et al (2020). Statistical significance of each TE estimate was evaluated using *F*-statistics based on the entropy terms, enabling inference on the presence and strength of directed physiological coupling across neural, cardiac, and respiratory systems (Morales et al., 2020).

### Statistical Analyses

In order to evaluate the effects of group (novices vs experts) and condition (meditation vs rest) on cardiorespiratory measures, encompassing respiration frequency, mean heart rate, and the respiration-adjusted HRV metric (Px), two-way mixed-design ANOVAs were used in R, with group as the between-subjects factor and condition as the within-subjects factor.

Regarding neurovisceral measures, to assess significant differences between conditions (meditation vs rest) and between groups (novices vs experts), cluster-based permutation statistical tests were applied to both the cross-frequency ratio distributions and transfer entropy (TE) measures. This non-parametric approach allowed for robust testing across electrodes while controlling for multiple comparisons. This approach, implemented via the FieldTrip MATLAB toolbox (Maris & Oostenveld, 2007), controls for type I error inflation associated with multiple comparisons across electrodes by leveraging a non-parametric Monte Carlo randomization framework. For within-group comparisons (e.g., meditation vs rest), paired-sample t-tests (*ft_statfun_depsamplesT*) were performed at each electrode for the variable of interest (either cross-frequency ratio percentages or TE values). Clusters were defined based on spatial adjacency of electrodes showing consistent effect direction (same sign of *t*-values), with permutation testing (10,000 iterations) used to generate a null distribution of cluster-level statistics (sum of *t*-values within each cluster). The cluster-corrected *p*-value was computed as the proportion of random permutations where the maximum cluster statistic exceeded that observed in the real data. The significance and cluster-defining thresholds were set at *p* < 0.05. For between-group comparisons (novices vs experts, at both rest and meditation), independent-sample *t*-tests were used (*ft_statfun_indepsamplesT*), again clustering across adjacent electrodes and performing the same permutation-based correction for multiple comparisons. This ensured that any group-related differences in ratio distributions or TE measures were robustly identified while controlling for the family-wise error rate across electrodes. In all analyses, electrodes were treated as the clustering dimension to capture spatially coherent effects, and identical statistical procedures were applied to both the cross-frequency ratio data and the four TE measures.

In order to explore potential group- and condition-related differences in cross-frequency ratios, analyses were first focused solely on the targeted ratio 8:1, selected based on prior work as a marker of potential alpha: heart rate coupling (Klimesch, 2018; Soriano et al., 2024). Subsequently, the analysis was extended to include all computed ratios to identify additional patterns in the cross-frequency relationships between alpha and heart rate.

### Additional analyses

#### Transfer entropy asymmetry

To further explore the directional dynamics between neural and physiological signals, asymmetry indices reflecting the relative strength of influence in each direction for both alpha–HRV and alpha–respiration interactions were calculated. For each participant, condition, and electrode, the asymmetry was computed by subtracting the TE value in one direction from the TE in the opposite direction (e.g., TE alpha→ HRV minus TE HRV→ alpha; and likewise, for alpha→ respiration vs respiration→ alpha). These asymmetry indices provided a direct measure of the dominant directionality of information flow within each system. To assess statistically significant differences in asymmetry between conditions (meditation vs rest) and between groups (novices vs experts), the same cluster-based permutation framework described above was applied.

#### Relationship between condition-related changes in physiological variables, TE and cross-frequency ratios

In order to obtain a more comprehensive assessment of the overall systemic dynamics, Pearson correlations were calculated between condition-related changes at the individual physiological measures (respiration rate, heart rate and Px), and condition-related changes in TE values in significant electrodes and their corresponding electrodes in cross-frequency ratios. Condition-related changes were calculated by subtracting each participant’s average values during meditation from values during the rest period. Correlation results were adjusted using false discovery rate (FDR) correction.

## Results

### Respiration rate group and condition effects

As a first step towards characterizing physiological differences between groups and conditions, respiration rate was examined across conditions of rest and meditation in both the novice and expert groups. Respiration rates exhibited a consistent linear decrease across conditions and groups (**Figure 1**). Mixed effect ANOVA analyses revealed a significant main effect of condition [*F*(1, 107) = 26.94, *p* < 0.001)], with participants showing higher respiration rates during rest (mean novices = 0.21 Hz; *SD* = 0.07; mean experts = 0.17 Hz; *SD* = 0.07) compared to meditation practice (mean novices = 0.16 Hz; *SD* = 0.07; mean experts = 0.13 Hz; *SD* = 0.06). A significant main effect of group was also found [*F*(1, 107) = 5.37, *p* = 0.02], revealing that experts had slower respiration rates than novices across conditions. No significant interaction effect was found between the two factors [*F*(1, 107) = 0.71, *p* = 0.4]. These findings confirmed the expected respiration reductions during meditation and further suggest a trait difference between novice and experienced meditators. However, pairwise comparisons exploring the difference between groups during either rest or meditation separately did not yield significant differences (all *p* > 0.05).

**Figure 1.**
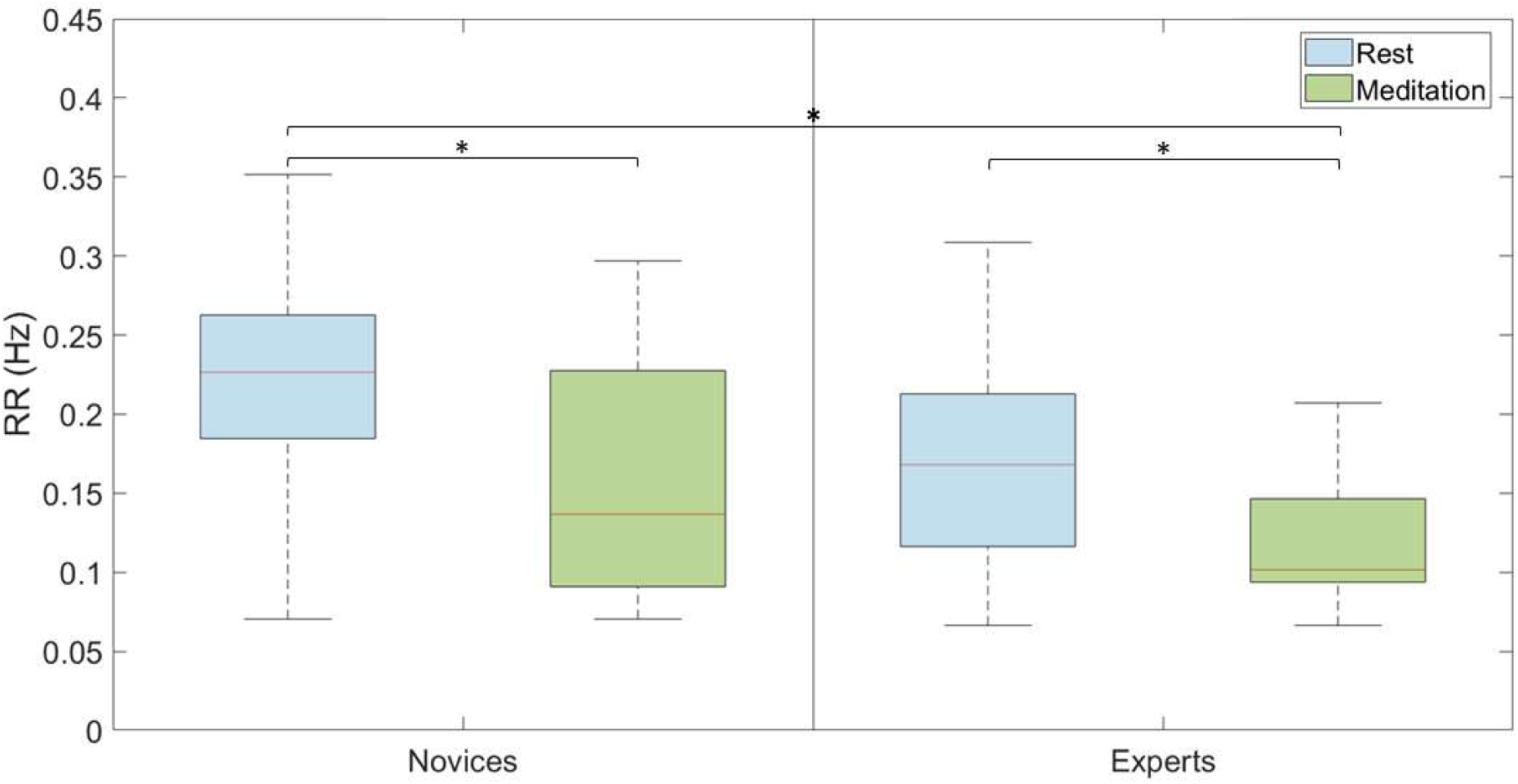
Condition and group-related differences in respiration rate. For each condition and group, box plots are presented, depicting median (red line), minimum and maximum values (bottom and top whiskers) as well as first (bottom) and third quartiles (top box boundary) of the respiration rate (RR), separately for the novices’ group during rest (yellow), novices during meditation (green), experts’ group during rest (blue) and experts during meditation (red). Significant differences (*p* <0.05) are indicated with an asterisk *.

### Heart rate group and condition effects

In contrast to respiration, heart rate did not differ significantly across groups or conditions (**Figure 2**), as revealed by mixed effect ANOVA analyses on the main factor condition [*F*(1, 107) = 1.26, *p* = 0.34)], group [*F*(1, 107) = 0.01, *p* = 0.92)] and the interaction thereof [*F*(1, 107) = 4.55, *p* = 0.05)]. Heart rate values were highly comparable across groups during both rest (mean novices = 1.13 Hz; *SD* = 0.13; mean experts = 1.15 Hz; *SD* = 1.17) and meditation (mean novices = 1.15 Hz; *SD* = 0.14; mean experts = 1.15 Hz; *SD* = 1.16), revealing that this cardiac measure remains unaffected by meditative practice and level of experience.

**Figure 2.**
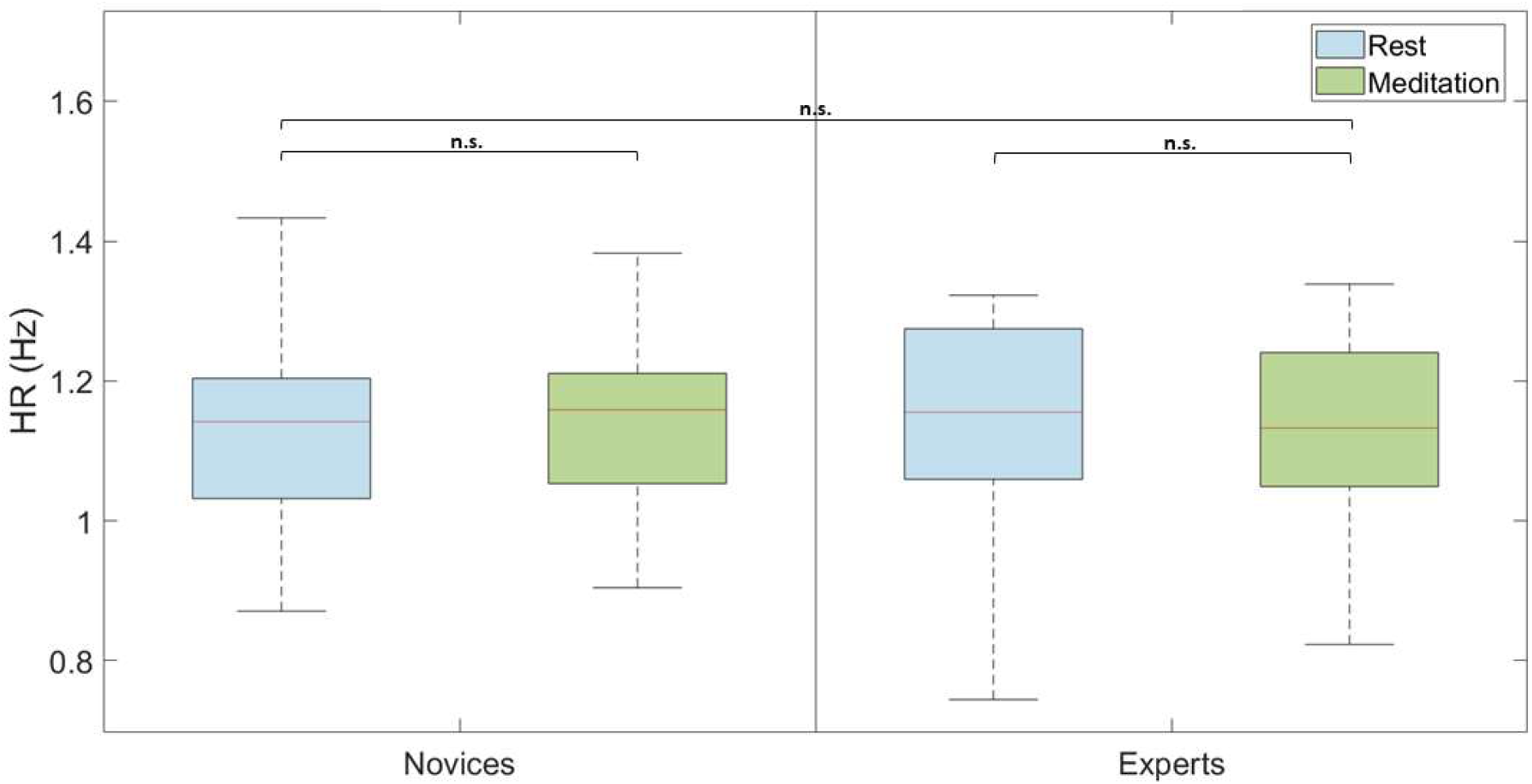
Condition and group-related differences in heart rate. For each condition and group, box plots are presented, depicting median (red line), minimum and maximum values (bottom and top whiskers) as well as first (bottom) and third quartiles (top box boundary) of the average heart rate (HR), separately for the novices’ group during rest (yellow), novices during meditation (green), experts’ group during rest (blue), and experts during meditation (red). Significant differences (*p* <0.05) are indicated with an asterisk *.

**Figure 3.**
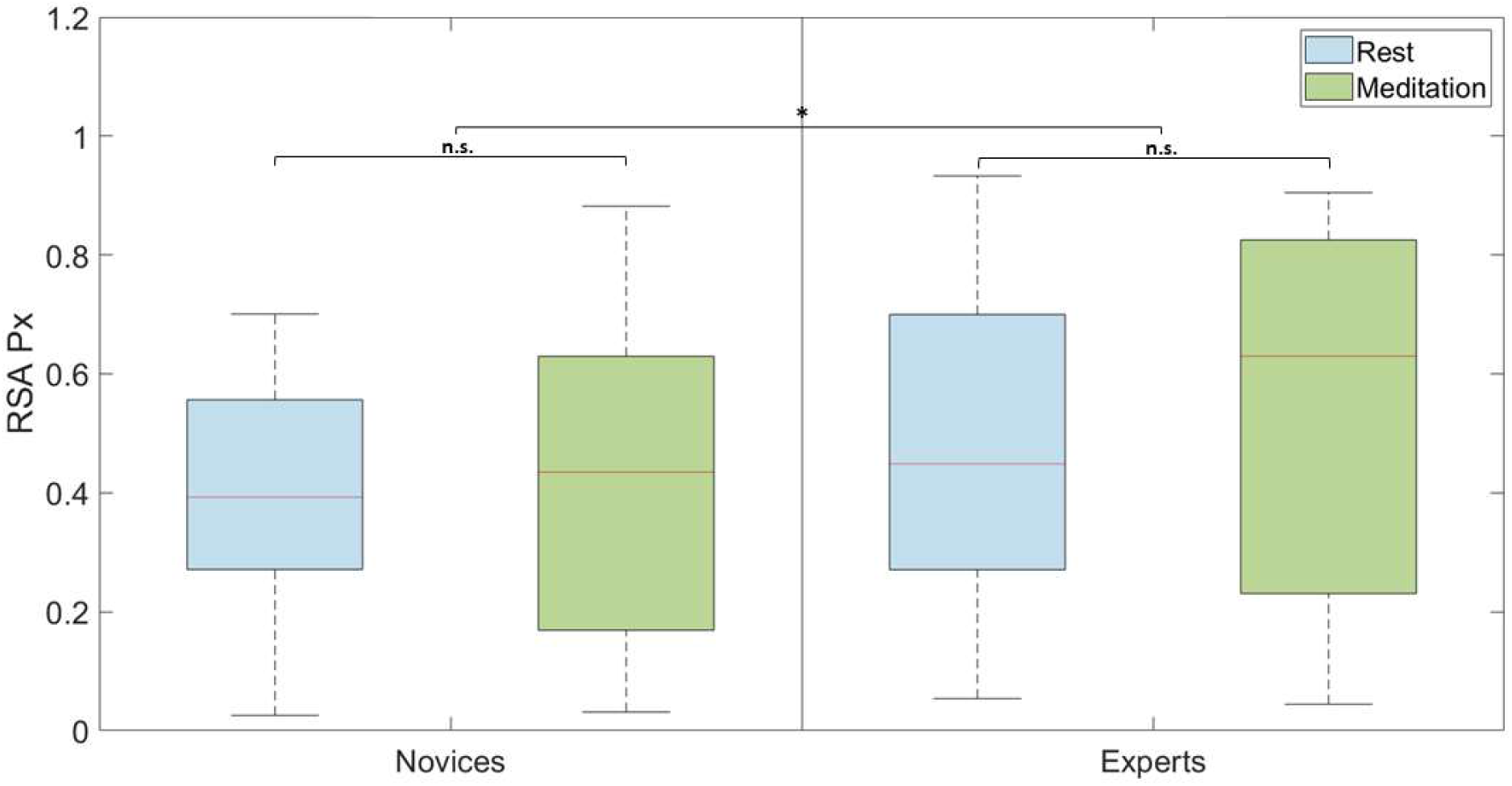
Group-related differences in RSA Px. For each group, box plots are presented, depicting median (red line), minimum and maximum values (bottom and top whiskers) as well as first (bottom) and third quartiles (top box boundary) of the average RSA Px index, separately for the novices (green) and experts (blue). Significant differences (*p* <0.05) are indicated with an asterisk *.

### Cardiorespiratory dynamics group and condition effects

To further characterize cardiorespiratory dynamics beyond heart rate, respiratory sinus arrhythmia using the Px index was examined (see Methods section). The mixed-effect model revealed a significant effect of group [*F*(1, 107) = 3.51, *p* < 0.05)], indicating greater parasympathetic influence in the expert meditator group (mean experts = 0.51; *SD* = 0.27) compared to the novices (mean novices = 0.41; *SD* = 0.23) across conditions. Conversely, neither the condition effect [*F*(1, 107) = 0.82, *p* = 0.37)] nor the interaction term reached significance [*F*(1, 107) = 0.32, *p* = 0.58)]. These findings suggest that expert meditators exhibit a state-independent elevated parasympathetic tone indicating a possible trait characteristic associated to their meditation experience.

### Neurocardiac Information Dynamics

#### Alpha to Heart Transfer Entropy group and condition effects

To explore functional coupling between cardiac and neural signals, directed information flow was examined through transfer entropy between alpha and cardiac activity. As depicted in **Figure 4**, cluster-based permutation analyses revealed no significant differences in transfer entropy from alpha to heart between novices and experts (**Figure 4F** and **H**) or within groups comparing meditation and rest conditions (**Figure 4E** and **G**). Thus, the influence of alpha activity over cardiac functioning remained stable regardless the task or meditation expertise.

**Figure 4.**
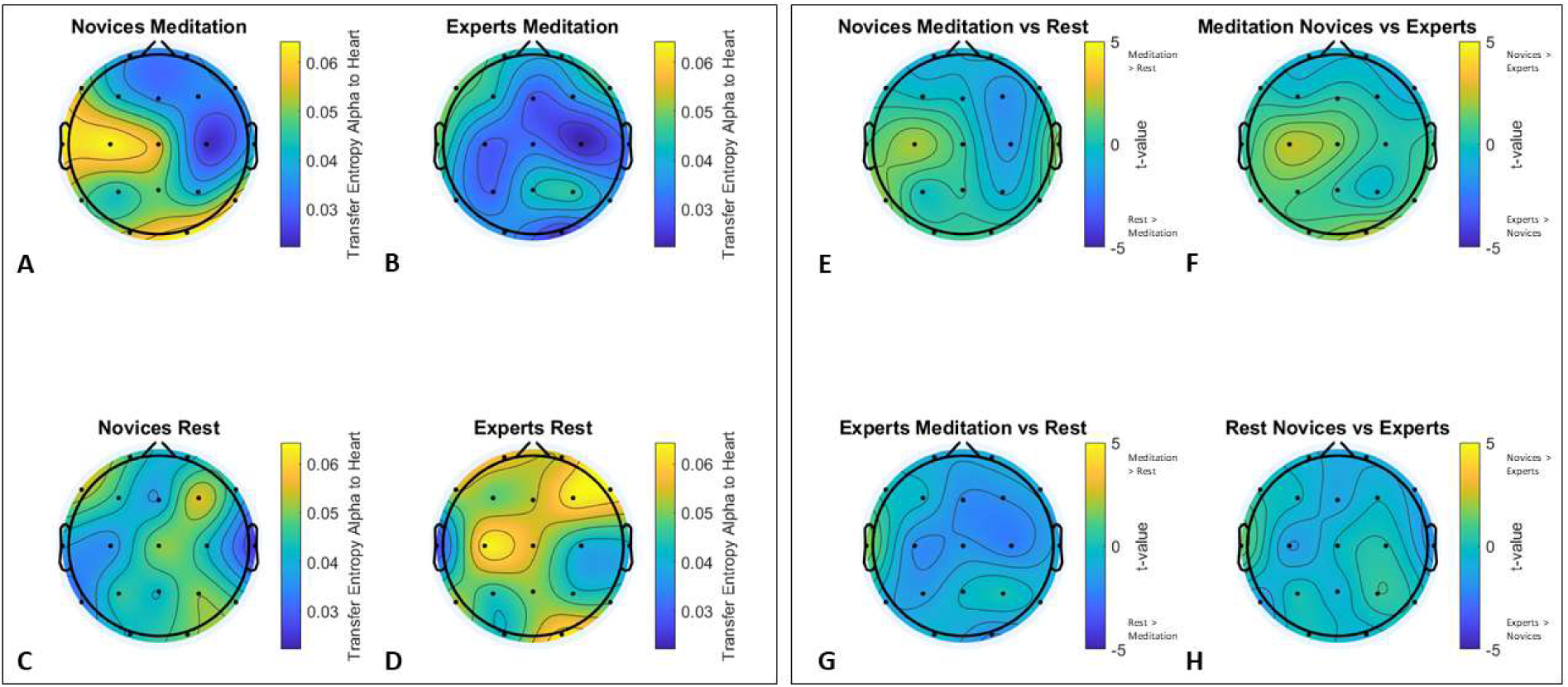
Transfer Entropies Alpha to Heart and permutation tests of the group and condition differences. **A-D)** Topographical plots depicting distributions of Transfer Entropies across groups and conditions. **E-H)** Topographical plots depicting differences between and within groups. Colourmaps code for the direction of the difference as represented by t-values. Asterisks indicate *p* < 0.05.

#### Heart to Alpha Transfer Entropy group and condition effects

Regarding differences in transfer entropy from cardiac activity to alpha (**Figure 5**), permutation tests yielded a cluster indicating significantly greater transfer entropy at electrode O2 during meditation in novices compared to experts (*t_cluster_* (19) = 2.93; *p* = 0.04). No other comparisons yielded significant differences. Although spatially restricted to a unilateral occipital region, this result indicates higher levels of influence of cardiac activity over alpha dynamics for the novices group compared to experts during meditation.

**Figure 5.**
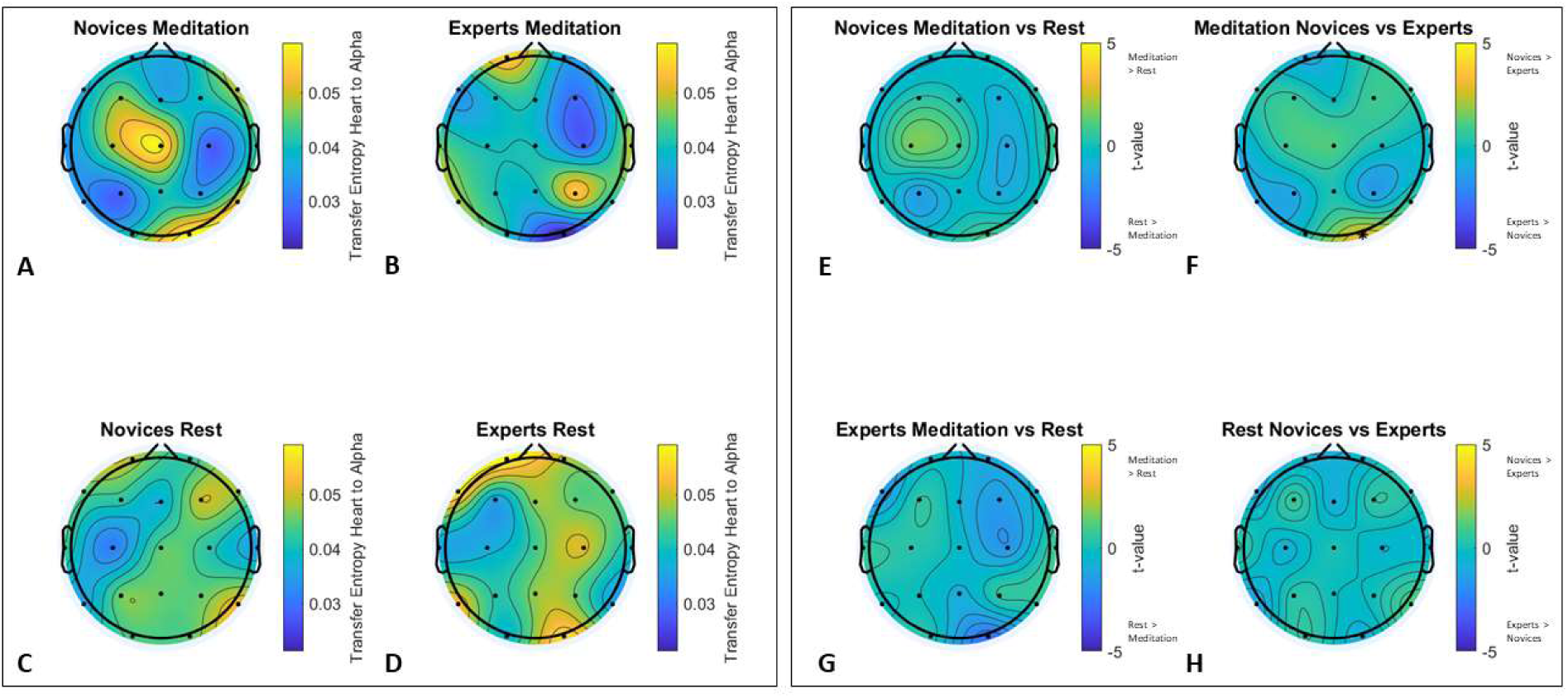
Transfer Entropies Heart to Alpha and permutation tests of the group and condition differences. **A-D)** Topographical plots depicting distributions of Transfer Entropies across groups and conditions. **E-H)** Topographical plots depicting differences between and within groups. Colourmaps code for the direction of the difference as represented by t-values. Asterisks indicate *p* < 0.05.

### Neurorespiratory Information Dynamics

#### Alpha to Respiration Transfer Entropy group and condition effects

Next, bidirectional coupling between alpha and respiratory activity was investigated. Regarding information flow from alpha to respiration (**Figure 6**), experienced meditators exhibited significantly greater transfer entropy during meditation compared to rest (**Figure 6G**), with a cluster observed at right prefrontal electrodes F4 and F8 (*t_cluster_* (19) = 4.48; *p* = 0.03). Additionally, during meditation, experts also showed higher transfer entropy than novices (**Figure 6F**) at frontocentral midline electrodes Fz and Cz (*t_cluster_* (19) = -4.36; *p* = 0.03). Overall, these results suggest that expertise in meditation enhances top-down neural modulation of respiratory activity whereas this modulation is not maintained as a trait during rest.

**Figure 6.**
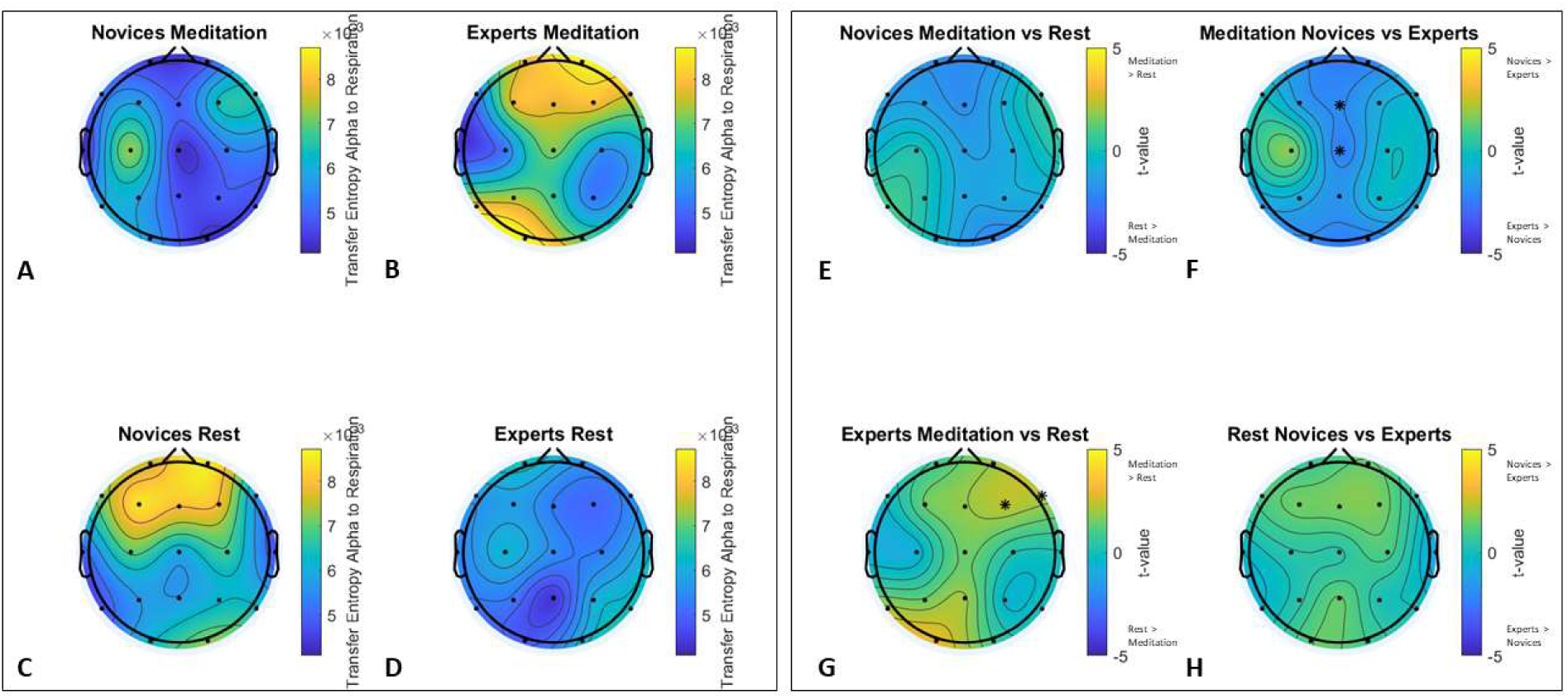
Transfer Entropies Alpha to Respiration and permutation tests of the group and condition differences. **A-D)** Topographical plots depicting distributions of Transfer Entropies across groups and conditions. **E-H)** Topographical plots depicting differences between and within groups. Colourmaps code for the direction of the difference as represented by t-values. Asterisks indicate *p* < 0.05.

#### Respiration to Alpha Transfer Entropy group and condition effects

In the case of information flow from respiration to alpha (**Figure 7**), cluster-based permutation tests yielded a cluster indicating significantly higher transfer entropy at electrode P4 in novices during meditation compared to rest (*t_cluster_* (19) = 2.78; *p* = 0.04). No other significant differences were found in this direction.

**Figure 7.**
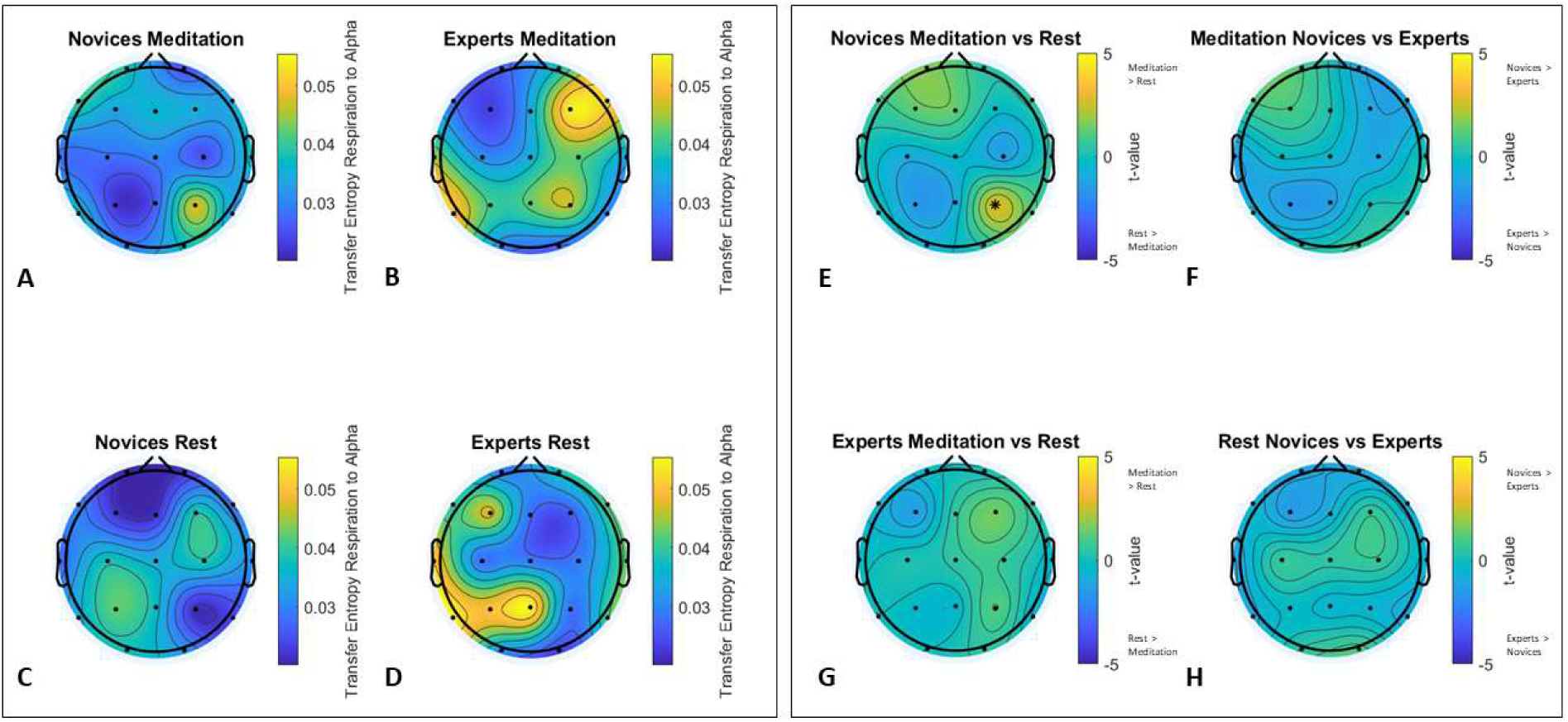
Transfer Entropies Respiration to Alpha and permutation tests of the group and condition differences. **A-D)** Topographical plots depicting distributions of Transfer Entropies across groups and conditions. **E-H)** Topographical plots depicting differences between and within groups. Colourmaps code for the direction of the difference as represented by t-values. Asterisks indicate *p* < 0.05.

#### Information flow asymmetry between respiration and alpha

Notably, across groups and conditions, it was observed that the magnitude of transfer entropy from respiration to alpha was consistently higher (values in the order of 10^-2^) compared to values of transfer entropy from alpha to respiration (values in the order of 10^-3^), suggesting a general asymmetry in information flow between the two physiological systems (see **Figure 5A-D** vs **6A-D**). To test for directional influence predominances, asymmetry scores were computed by subtracting alpha-to-respiration transfer entropy from respiration-to-alpha transfer entropy per participant and condition. Cluster-based permutation tests on these scores revealed that only in the case of novices, when comparing information flow asymmetry between meditation and rest, there was a significant predominance of information flow from respiration to alpha at electrode P4 (*t_cluster_* (19) = -2.98; *p* = 0.03), consistent with the results shown in Figure 7. Comparisons among experts and between groups in the same condition (meditation and rest) resulted in no significant findings (all *p* > 0.05).

Overall, the observed patterns in neurovisceral coupling pairs reveal a state and experience-dependent modulation. Whereas novices seem to rely on bottom-up influences from respiratory activity during meditation compared to rest, and influences from cardiac activity during meditation when compared to experts, expert meditators engage top-down neural to respiratory regulation processes.

#### Condition-related differences in transient alpha: heart rate cross-frequency ratios

Similar to previous work (Soriano et al., 2024), putative condition-related modulations in instantaneous alpha: heart rate cross-frequency ratios between groups and conditions were assessed. In **Figure 8**, the percentage (proportion of epochs) of each possible ratio is depicted for each electrode location. For each condition and group, cross-frequency ratios followed a normal distribution, similar across the entire scalp, with a maximal incidence at ratio 8 for the experts (13.04 %) and 8.5 for the novices (13.23 %) during the rest condition, and also at ratio 8 for the experts (10.66 %) and ratio 9 for the novices (12.98 %) during the meditation condition.

**Figure 8.**
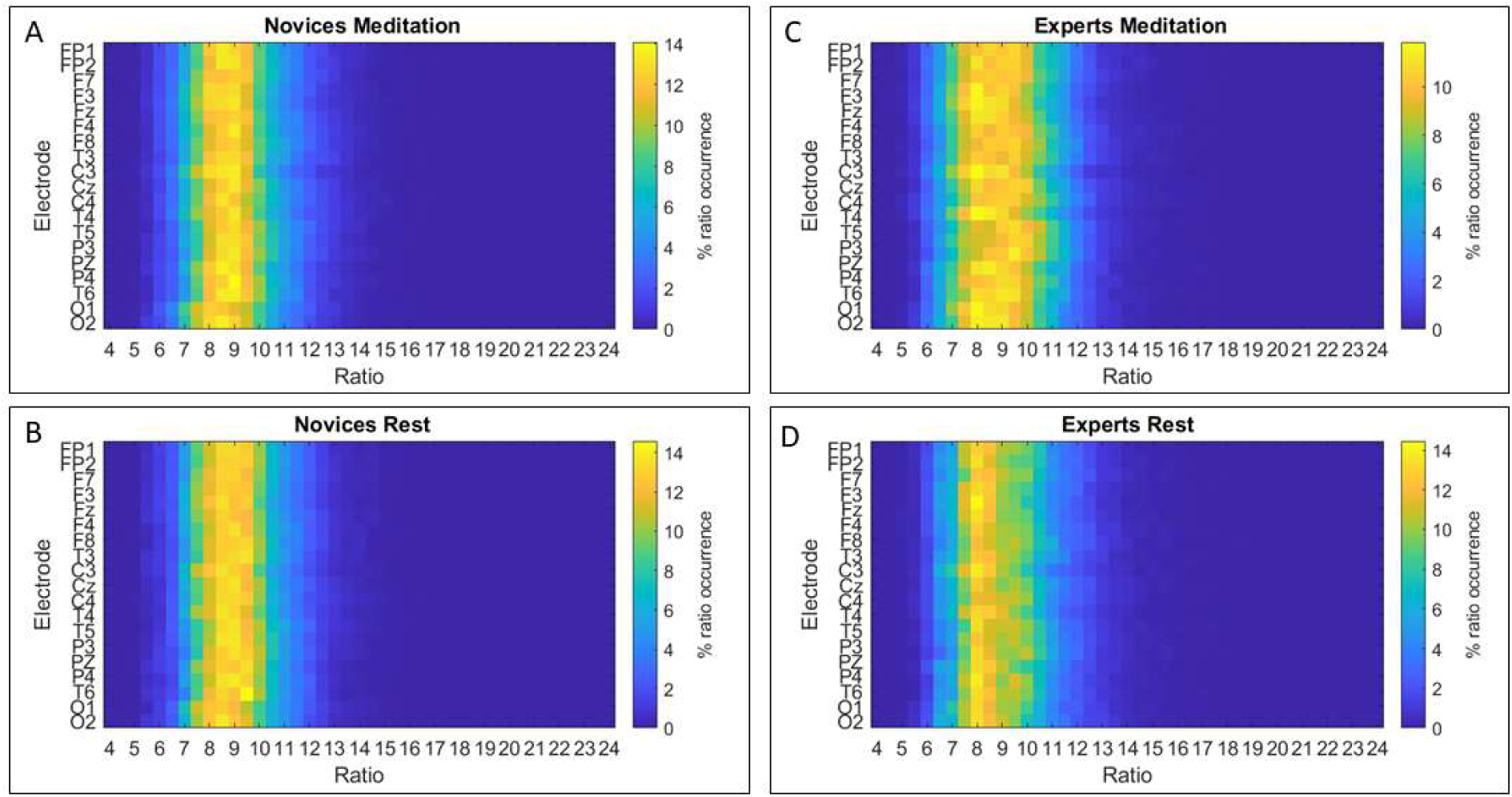
Distribution of the incidence (% of epochs) of alpha: heart rate cross-frequency ratios. For each electrode (y-axis) and cross-frequency ratio (x-axis) the percentage of incidence is shown according to the colormap in A) for the novices during meditation, B) novices during rest, C) experts during meditation and D) experts during rest.

Building on prior findings of condition-related differences in the specific harmonic 8:1 ratio (Soriano et al., 2024), a targeted hypothesis-driven analysis revealed that, in expert meditators, engagement in the meditation condition ,as compared to rest, was associated with a significant reduction in the incidence of the harmonic 8:1 alpha–heart rate ratio at electrodes, P3 (*t_cluster_* (19) = -2.81; *p* = 0.04) and Pz (*t_cluster_* (19) = -2.57; *p* = 0.04) . In contrast, no condition-related differences in this ratio were observed among novices, nor were there any direct group-related differences between novices and experts.

Next, and similar to previous work (Soriano et al., 2024), cluster-based permutation statistics were conducted to examine condition-related differences across all cross-frequency ratios. Despite observable trends as shown in **Figure 9**, no effects were significant (all *p*-values associated to the presented *t*-values were > 0.05). Overall, the lack of group and condition effects is likely due to the absence of heart rate differences, as demonstrated in prior work (Soriano et al., 2024).

**Figure 9.**
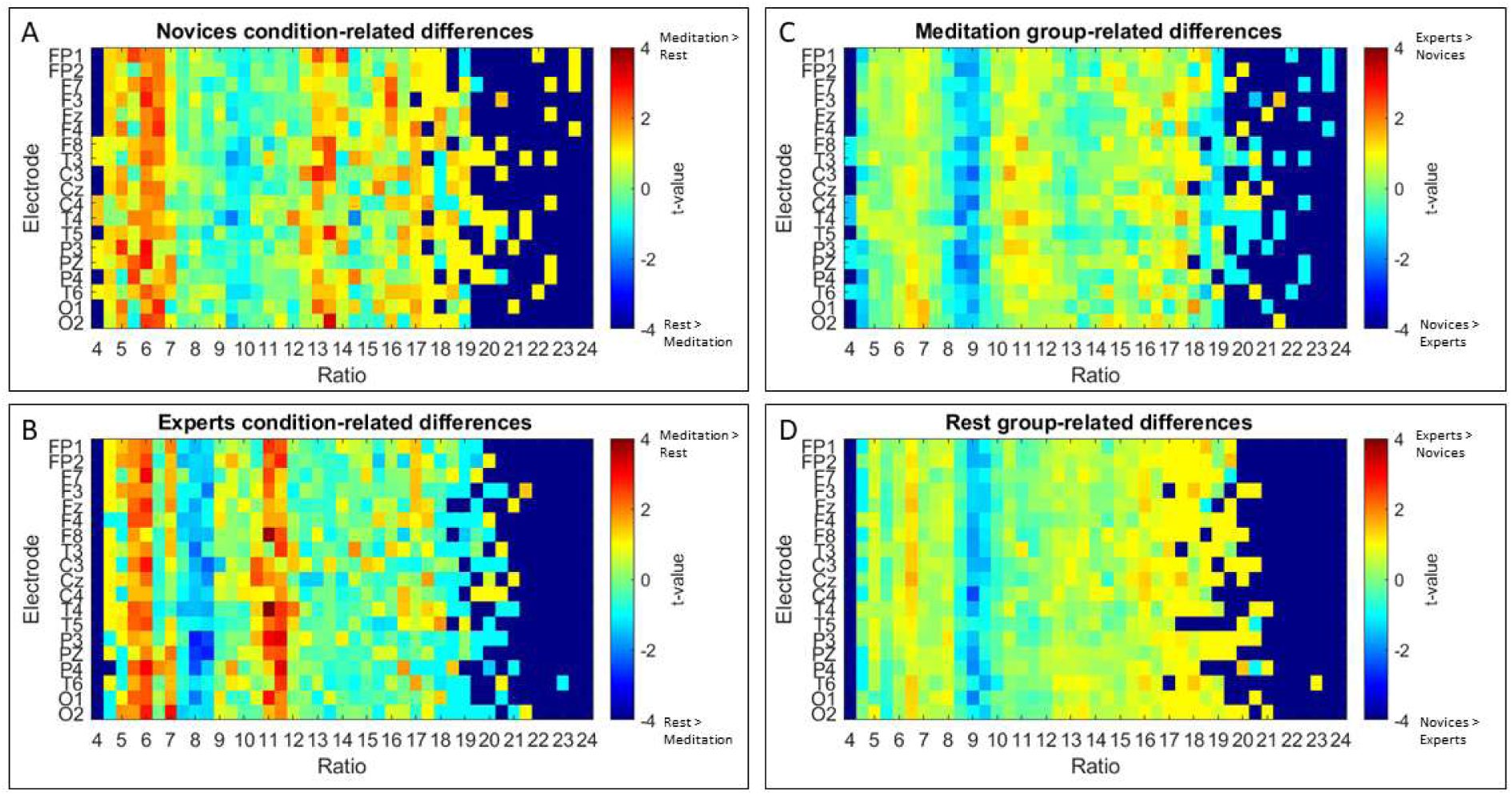
Condition- and group-related differences in the incidence (% of epochs) of different alpha: heart rate cross-frequency ratios. A) visualizes the colormap of t-values derived from cluster-based permutation analyses estimating the condition-related effect in novices (paired-sample ttest; meditation versus rest condition), separately for each electrode (y-axis) and cross-frequency ratio (x-axis). B) shows the map of t-values for the condition-related effect in experts. C) and D) show the t-values for the group-related effect during meditation and rest respectively.

#### Correlation analyses between transfer entropy, cross-frequency relationships and physiology

Considering condition and group related differences in cardiorespiratory measures, neurovisceral transfer entropies and the cross-frequency ratio 8:1, correlations were explored amongst these measures at scalp locations derived from the previous analyses. No correlations reached statistical significance (all *p* > 0.05).

## Discussion

This study investigated cardiorespiratory and neurovisceral integration dynamics during a period of breath-focused meditation and at rest, in both novice and expert meditators. While heart rate remained stable across conditions and groups, meditation was associated with reduced respiratory rates, particularly in expert practitioners, who also showed higher parasympathetic activity across conditions. Directional information flow analyses revealed that novices exhibited stronger bottom-up influences from cardiac to alpha activity during meditation compared to experts and from respiration to alpha compared to rest, both effects particularly at posterior sites. In contrast, experts showed enhanced top-down alpha-to-respiration modulation during meditation, particularly at frontocentral midline and right prefrontal regions. In order to investigate potential mechanisms for the observed bidirectional influences, targeted hypothesis-driven cross-frequency ratio analyses were used (Klimesch, 2018), revealing a reduced occurrence of the ratio 8:1 when experts engaged in the meditation condition as compared to rest. Cluster-based analyses encompassing all ratios, however, did not reveal any significant differences across groups or conditions.

### Respiratory, cardiac and cardiorespiratory effects

Regarding respiratory activity changes, results are in line with previous literature showing a decrease in respiratory rate during mindfulness meditation compared to rest and other control conditions (Ahani et al., 2014; Atchley et al., 2016) and at rest after mindfulness practices (Dietrich & Bidart, 2024; Karunarathne et al., 2024; Rusinova et al., 2024; Wielgosz et al., 2016). Furthermore, the present study supports the hypothesis that such reductions are not restricted to the meditation practice, but extend to reflect a trait observable in expert meditators, also consistent with previous reports (Gerritsen & Band, 2018; Wielgosz et al., 2016). Importantly, meditation practices such as focused attention on the breath do not necessarily instruct altering breathing patterns, distinguishing dhyāna/awareness from pranayama/breath control techniques (Balban et al., 2023; Bentley et al., 2023). The findings about deceleration of respiratory rate here may reflect both intentional regulation of the depth and pace of inhalations and exhalations, as well as a reduced metabolic demand associated with the relaxed state induced by breath-focused practices (Zaccaro et al., 2018). Alternatively, these reductions may arise from an ideomotor effect, where focused attention on breathing unconsciously influences respiratory patterns without explicit effort, or from the integration of complementary practices, particularly among expert meditators, where a persistently lower respiratory rate has been observed as a trait characteristic in the present and other studies (Gerritsen & Band, 2018; Wielgosz et al., 2016).

To further investigate the physiological underpinnings of breath-focused meditation practice and expertise, and to test for potential effects of relaxation in this specific practice, cardiac and cardiorespiratory activities were also investigated. Similar to previous work (Dietrich & Bidart, 2024; Guo et al., 2022; Jiang et al., 2020; Soriano et al., 2024), no significant differences in heart rate across groups or conditions were found. Although heart rate has been shown to change significantly when comparing stressful tasks to meditative practices (Soriano et al., 2024; Sun et al., 2019), results in this study mount to the evidence that this basic cardiac functioning marker is not able to capture subtle autonomic changes between two non-stressful states, such as rest and a breath-focused meditation practice. Heart rate variability (HRV), in contrast, encompasses a widely validated set of parameters that better reflects autonomic regulation, and inform on the fluctuations in sympathetic and parasympathetic activity, making it a more reliable indicator of relaxation than HR alone (Shaffer & Ginsberg, 2017). Crucially, the present study addresses a long-standing methodological limitation encountered by most studies assessing frequency domain HRV in the presence of slow respiratory rates, whereby conclusions on parasympathetic activity through the interpretation of typical HF HRV are confounded (Laborde et al., 2017; Quintana & Heathers, 2014; Ritz, 2024a, 2024b; Shaffer & Ginsberg, 2017; Varon et al., 2019). Here, greater parasympathetic tone was observed in expert meditators regardless of the condition, unparalleled by any heart rate modulations, and controlled for the occurrence of slower respiration rates. The presence of a group difference but absence of a condition effect indicates a trait effect consistent with the notion that meditation training derives in autonomic flexibility improvements in experts yet not observed in novices (Brown et al., 2021; Gerritsen & Band, 2018).

### Neurocardiac and neurorespiratory effects

In addition to parasympathetic regulation differences observed through cardiorespiratory measures, distinct patterns of neurovisceral influences during meditation were identified, with directionality (top-down vs. bottom-up) varying by expertise. Specifically, increased transfer entropy from cortical alpha to respiratory activity in expert meditators, but not in novices, was observed. Importantly, this effect was localized at electrode sites F4 and F8, partially coincident with areas such as the dorsolateral prefrontal cortex (DLPFC) and the inferior frontal gyrus (IFG), involved in top-down interoceptive mechanisms as well as emotional regulation (Sugawara et al., 2024) and somatic-motor processes. An increase in (top-down) alpha activity in the DLPFC area may reflect enhanced suppression of potentially distracting stimuli during interoceptive tasks such as focusing on the breath (Jelinčić et al., 2022), where greater activity during meditation in experts is expected due to their long-term training in the skill of letting distracting thoughts pass by, while redirecting the attention to the breath.

Additionally, midline electrodes Fz and Cz, positioned over the dorsal anterior cingulate cortex (dACC), supplementary motor area (SMA), and primary sensorimotor cortices, exhibited greater alpha to respiration transfer entropy in expert meditators compared to novices during meditation. This finding suggests that expert practitioners engage more selective interoceptive monitoring and motor-autonomic synchronization during practice, consistent with their greater capacity to tune in to body sensations and ignore task-irrelevant stimuli. Opposite to the pattern of frontocentral midline top-down modulation in experts, novices displayed a localized posterior (coincident with sensory-dominant areas), bottom-up directionality from both the cardiac and the respiratory domains, suggesting higher neural reliance and reactivity to visceral stimuli. More specifically, this effect was present for respiratory influences during breath-focused meditation relative to rest, and for cardiac influences during meditation as compared to experts.

Although this study is, to our knowledge, the first to investigate novice and expert meditators during both rest and meditation practice, both from a neurocardiac and a neurorespiratory perspective, previous studies have investigated a set of these groups, conditions and measures. For example, in a study by Wang et al., (2024), experts and novices were assessed during rest, and an increased correlation between fm-theta activity with RMSSD was found in experts, indicating enhanced top-down parasympathetic regulation. While using different HRV and neural measures, a similar pattern of enhanced top-down regulation in expert meditators was found here, although restricted to respiratory activity, and also reflected in heightened parasympathetic tone. Along the same line, previous fMRI research has showed greater activation in both the DLPFC and the ACC in expert compared to non-expert meditators during breath-focused meditation practices (Short et al., 2010), largely consistent with other literature examining these areas during focused attention meditation (see Fox et al., 2016; Ganesan et al., 2022; Zsadanyi et al., 2021). It is likely that, as opposed to Wang et al., (2024), the lack of significant linear relationships between transfer entropy and cardiorespiratory measures in the present work stems from the presence of non-linear instead of linear relationships, which were already captured to an extent by the transfer entropy measures results.

### A predictive processing perspective

From a theoretical standpoint, the predictive processing framework offers a compelling lens through which to interpret the present results (Braendholt et al., 2023). This framework proposes that the brain continuously generates and updates internal models to anticipate sensory input, minimizing prediction errors through a balance of top-down predictions and bottom-up signals (Friston, 2010). Within this model, the enhanced top-down alpha to respiration coupling found in experts could be interpreted as an increased precision-weighting of interoceptive predictions selective to respiration while suppressing irrelevant noise, which further impacts respiratory activity to effectively fit such predictions (Friston, 2019). Indeed, the localization of the effects over frontocentral midline and right prefrontal regions is consistent with predictive coding models stablishing a control hierarchy initiated in these areas (Friston, 2019). In contrast, the effects found on novices manifesting more posterior bottom-up responses, point to a greater dependence on sensory input, reflecting earlier stages of interoceptive skill development where prediction models are less refined due to lack of experience. These converging results suggest the notion that meditation training might foster neurovisceral integration by progressively shifting the balance from reactive (bottom-up) to proactive (top-down) interoceptive control, along the broader process of adaptive predictive processing.

### Effects on cross-frequency relationships

In addition to the exploration of bidirectional neurovisceral influences through the information dynamics framework, and similar to previous work (Soriano et al., 2024), the cross-frequency ratio approach proposed by Klimesch (2018) was also applied, in order to test for underlying mechanisms of neurocardiac interactions in terms of cross-frequency relationships. Interestingly, the overall incidence of the harmonic 8:1 alpha: heart rate ratio was found to be maximal in experts across conditions. Further targeted analyses showed that, in a comparison between meditation and rest within the expert meditator group, this ratio was more prominent during rest. This finding extends earlier observations showing that, during an arithmetic task, the 8:1 cross-frequency relationship is more frequently expressed, presumably facilitating neural–cardiac coupling during heightened cognitive effort (Soriano et al., 2024). In the current study, the reduced incidence of this ratio during meditation among expert meditators, may similarly reflect a shift away from such effortful coupling, possibly indicating a state of reduced cognitive demand or greater autonomic regulation associated with meditative practice. However, caution should be taken with respect to interpreting these findings, since whole-scalp exploratory analyses across all cross-frequency ratios failed to yield any significant cluster when adopting robust statistical measures for adjusting multiple testing. In light of earlier findings in novices, several methodological differences likely explain the less robust findings regarding differences in the ratio 8∶1. In the previous study including only novices (Soriano et al., 2024), the design included a meditation period and an arithmetic task, during which pronounced HR accelerations served as a primary driver of the found differences in the 8:1 cross-frequency ratio. By contrast, neither expert not novice meditators in the current study exhibit HR shifts across meditation and rest. Future investigations should therefore include parallel arithmetic or other demanding tasks in both novice and expert cohorts to disentangle the contributions of HR dynamics versus intrinsic neurovisceral coupling to cross-frequency effects.

In order to identify co-occurrences between single physiological and neurovisceral changes across groups and conditions, correlations were computed amongst the mentioned measures. No significant correlations were found, further reflecting a lack of redundancy between the used metrics and highlighting the need for a complementary and comprehensive assessment of physiological systems and their interdependence.

### Limitations and Future Directions

Although the present study offers novel insights into neurovisceral integration during breath-focused meditation, several limitations should be noted. A primary limitation is the use of a low-density EEG montage, which restricted the ability to perform precise source localization of the transfer entropy effects. Although the observed patterns over frontocentral midline and right prefrontal electrodes are consistent with prior research implicating the DLPFC, ACC, and IFG in interoceptive and autonomic control, higher-density EEG or MEG would enable a more fine-grained identification of the neural underpinnings of the observed directional information flow patterns.

While transfer entropy effectively captures directed functional connectivity, it does not distinguish between volitional control and more automatic or ideomotor processes. This is particularly relevant for interpreting the cognitive process underlying top-down alpha to respiration modulations, where it remains unclear whether participants were actively regulating their breath or whether attentional focus alone elicited unconscious adjustments. To address this, future studies should incorporate subjective measures (e.g., experience sampling) to assess the degree of perceived effort, agency, and awareness regarding breath control during meditation.

Another important limitation concerns the treatment of the respiratory signal regardless its phase. Although respiratory rates and modulations were analyzed globally, growing evidence underscores that inhalation and exhalation phases are functionally distinct and may differentially couple with and entrain neural oscillations (Herrero et al., 2018; Zaccaro et al., 2025). Future work should warrant phase-specific analyses, such as examining phase-amplitude coupling between inhalation/exhalation phases and cortical rhythms, to better disentangle the temporal dynamics of neuro-respiratory integration.

Regarding the experts’ sample, it is worth noting that the studied group encompassed certain variability in meditation experience (e.g., years of practice, retreat experience, type of training), which was not stratified in the analyses due to insufficient sample size. Future research could benefit from a more granular categorization of expertise levels and meditation styles to examine their potentially distinct contributions to neurovisceral integration. In sum, while this study contributes to growing evidence for distinct neurovisceral integration patterns shaped by meditation expertise, more nuanced methodological designs and mechanistic probes are needed to fully unravel the complex interplay between neural and visceral physiologies underpinning autonomic control and interoception in contemplative practices.

## Conclusion

This study offers a novel perspective on how contemplative practices modulate neurovisceral integration, advancing methodological and theoretical approaches for the investigation of multimodal systemic and complex dynamics. By integrating measures of autonomic regulation with directional information dynamics and cross-frequency neural analyses, the presented results reveal that experienced practitioners exhibit enhanced top–down cortical modulation of respiration alongside a marked increase of parasympathetic activity across both rest and meditation conditions. These findings highlight how breath-focused meditation can progressively reshape the dynamic interplay between attentional processes and autonomic regulation, offering insights for advancing the understanding of contemplative, interoceptive, and self-regulatory processes. More broadly, the contributions of this study provide additional evidence to advance investigations into embodied cognition, mental well-being, and the refinement of contemplative and therapeutic interventions, such as the development of combined bio- and neurofeedback protocols based on interactions between visceral and neural signals.

## Author Contribution

Javier R. Soriano: conceptualization, methodology, software, formal analysis, writing —original draft, visualization, supervision, project administration. Angeliki-Ilektra Karaiskou: software, formal analysis, writing review and editing. Julio Rodriguez-Larios: investigation, resources, data curation, writing — review and editing, funding acquisition. Carolina Varon: supervision, software, writing — review and editing. Nazareth Castellanos: supervision, writing — review and editing. Kaat Alaerts: conceptualization, writing — review and editing, supervision, funding acquisition.

## Data Availability

Raw data as well as MATLAB code will be made publicly available in the KU Leuven Research Data Repository upon request and/or after acceptance for publication.

## References

1. Adelhöfer, N., Schreiter, M. L., & Beste, C. (2020). Cardiac cycle gated cognitive-emotional control in superior frontal cortices. NeuroImage, 222, 117275. 10.1016/j.neuroimage.2020.117275

2. Ahani, A., Wahbeh, H., Nezamfar, H., Miller, M., Erdogmus, D., & Oken, B. (2014). Quantitative change of EEG and respiration signals during mindfulness meditation. Journal of NeuroEngineering and Rehabilitation, 11(1), 1–11. 10.1186/1743-0003-11-87

3. Ainsworth, B., Eddershaw, R., Meron, D., Baldwin, D. S., & Garner, M. (2013). The effect of focused attention and open monitoring meditation on attention network function in healthy volunteers. Psychiatry Research, 210(3), 1226–1231. 10.1016/j.psychres.2013.09.002

4. Allen, M., Varga, S., & Heck, D. H. (2022). Respiratory Rhythms of the Predictive Mind. Psychological Review, 130(4), 1066–1080. 10.1037/rev0000391

5. Anurag, S., Singh, B. K., Krishna, D., Prasanna, K., & Deepeshwar, S. (2023). Heart–brain Rhythmic Synchronization during Meditation: A Nonlinear Signal Analysis. International Journal of Yoga, 16(2), 132–139. 10.4103/ijoy.ijoy_161_23

6. Ashhad, S., Kam, K., Del Negro, C. A., & Feldman, J. L. (2022). Breathing Rhythm and Pattern and Their Influence on Emotion. Annual Review of Neuroscience, 45, 223–247. 10.1146/annurev-neuro-090121-014424

7. Atad, D. A., Mediano, P. A. M., Rosas, F. E., & Berkovich-ohana, A. (2023). Meditation and Complexity : a Systematic Review. 10.31234/osf.io/np97r

8. Atchley, R., Klee, D., Memmott, T., Goodrich, E., Wahbeh, H., & Oken, B. (2016). Event-related potential correlates of mindfulness meditation competence. Neuroscience, 320, 83–92. 10.1016/j.neuroscience.2016.01.051

9. Azzalini, D., Rebollo, I., & Tallon-Baudry, C. (2019). Visceral Signals Shape Brain Dynamics and Cognition. In Trends in Cognitive Sciences (Vol. 23, Issue 6, pp. 488–509). Elsevier Ltd. 10.1016/j.tics.2019.03.007

10. Bailey, N. W., Fulcher, B. D., Caldwell, B., Hill, A. T., Fitzgibbon, B., van Dijk, H., & Fitzgerald, P. B. (2024). Uncovering a stability signature of brain dynamics associated with meditation experience using massive time-series feature extraction. Neural Networks, 171(June 2023), 171–185. 10.1016/j.neunet.2023.12.007

11. Baird, B., Mrazek, M. D., Phillips, D. T., & Schooler, J. W. (2014). Domain-specific enhancement of metacognitive ability following meditation training. Journal of Experimental Psychology: General, 143(5), 1972–1979. 10.1037/a0036882

12. Balzarotti, S., Biassoni, F., Colombo, B., & Ciceri, M. R. (2017). Cardiac vagal control as a marker of emotion regulation in healthy adults: A review. Biological Psychology, 130(April), 54–66. 10.1016/j.biopsycho.2017.10.008

13. Baminiwatta, A., & Solangaarachchi, I. (2021). Trends and Developments in Mindfulness Research over 55 Years: A Bibliometric Analysis of Publications Indexed in Web of Science. Mindfulness, 12(9), 2099–2116. 10.1007/s12671-021-01681-x

14. Bartsch, R. P., Liu, K. K. L., Bashan, A., & Ivanov, P. C. (2015). Network Physiology: How Organ Systems Dynamically Interact. PLOS ONE, 10(11), e0142143. 10.1371/journal.pone.0142143

15. Belli, F., & Fischer, M. H. (2024). Breathing shifts visuo-spatial attention. Cognition, 243(December 2023), 105685. 10.1016/j.cognition.2023.105685

16. Bentley, T. G. K., D’Andrea-Penna, G., Rakic, M., Arce, N., LaFaille, M., Berman, R., Cooley, K., & Sprimont, P. (2023). Breathing Practices for Stress and Anxiety Reduction: Conceptual Framework of Implementation Guidelines Based on a Systematic Review of the Published Literature. Brain Sciences, 13(12). 10.3390/brainsci13121612

17. Braendholt, M., Kluger, D. S., Varga, S., Heck, D. H., Gross, J., & Allen, M. (2023). Breathing in waves: Understanding Respiratory-Brain Coupling as a Gradient of Predictive Oscillations. Neuroscience and Biobehavioral Reviews, 152(June), 105262. 10.1016/j.neubiorev.2023.105262

18. Brandmeyer, T., & Delorme, A. (2018). Reduced mind wandering in experienced meditators and associated EEG correlates. Experimental Brain Research, 236(9), 2519–2528. 10.1007/s00221-016-4811-5

19. Brandmeyer, T., Delorme, A., & Wahbeh, H. (2019). The neuroscience of meditation: classification, phenomenology, correlates, and mechanisms. Progress in Brain Research, 244, 1–29. 10.1016/bs.pbr.2018.10.020

20. Brown, L., Rando, A. A., Eichel, K., Van Dam, N. T., Celano, C. M., Huffman, J. C., & Morris, M. E. (2021). The Effects of Mindfulness and Meditation on Vagally Mediated Heart Rate Variability: A Meta-Analysis. Psychosomatic Medicine, 83(6), 631–640. 10.1097/PSY.0000000000000900

21. Candia-Rivera, D., Engelen, T., Babo-Rebelo, M., & Salamone, P. C. (2024). Interoception, network physiology and the emergence of bodily self-awareness. Neuroscience and Biobehavioral Reviews, 165(June), 0–2. 10.1016/j.neubiorev.2024.105864

22. Candia-Rivera, D., Norouzi, K., Ramsøy, T. Z., & Valenza, G. (2023). Dynamic fluctuations in ascending heart-to-brain communication under mental stress. American Journal of Physiology - Regulatory Integrative and Comparative Physiology, 324(4), R513–R525. 10.1152/ajpregu.00251.2022

23. Catrambone, V., Barbieri, R., Wendt, H., Abry, P., & Valenza, G. (2021). Functional brain-heart interplay extends to the multifractal domain. *Philosophical Transactions of the Royal Society A: Mathematical*, Physical and Engineering Sciences, 379(2212). 10.1098/rsta.2020.0260

24. Catrambone, V., Zallocco, L., Ramoretti, E., Mazzoni, M. R., Sebastiani, L., & Valenza, G. (2024). Integrative neuro-cardiovascular dynamics in response to test anxiety: A brain-heart axis study. Physiology and Behavior, 276(January), 114460. 10.1016/j.physbeh.2024.114460

25. Chan, R. W., Immink, M. A., & Lushington, K. (2017). The influence of focused-attention meditation states on the cognitive control of sequence learning. Consciousness and Cognition, 55(March), 11–25. 10.1016/j.concog.2017.07.004

26. Chapeton, J. I., Haque, R., Wittig, J. H., Inati, S. K., & Zaghloul, K. A. (2019). Large-Scale Communication in the Human Brain Is Rhythmically Modulated through Alpha Coherence. Current Biology, 29(17), 2801–2811.e5. 10.1016/j.cub.2019.07.014

27. Chems, R., Kate, M., Ruth, C., Jenny, B., & Clara, G. (2025). Defining Mindfulness : A Review of Existing Definitions and Suggested Refinements. Mindfulness. 10.1007/s12671-024-02507-2

28. Chiesa, A., & Serretti, A. (2009). Mindfulness-based stress reduction for stress management in healthy people: A review and meta-analysis. Journal of Alternative and Complementary Medicine, 15(5), 593–600. 10.1089/acm.2008.0495

29. Corcoran, A. W., Perrykkad, K., Feuerriegel, D., & Robinson, J. E. (2023). Body as First Teacher: The Role of Rhythmic Visceral Dynamics in Early Cognitive Development. Perspectives on Psychological Science. 10.1177/17456916231185343

30. Davies, J. N., Faschinger, A., Galante, J., & Van Dam, N. T. (2024). Prevalence and 20-year trends in meditation, yoga, guided imagery and progressive relaxation use among US adults from 2002 to 2022. Scientific Reports, 14(1), 1–11. 10.1038/s41598-024-64562-y

31. de Zambotti, M., Trinder, J., Silvani, A., Colrain, I. M., & Baker, F. C. (2018). Dynamic coupling between the central and autonomic nervous systems during sleep: A review. Neuroscience & Biobehavioral Reviews, 90(3), 84–103. 10.1016/j.neubiorev.2018.03.027

32. Delorme, A., & Makeig, S. (2004). EEGLAB: an open sorce toolbox for analysis of single-trail EEG dynamics including independent component anlaysis. Journal of Neuroscience Methods, 134, 9–21. 10.1016/j.jneumeth.2003.10.009

33. Dietrich, K. M., & Bidart, M. G. (2024). Effects of a mindfulness course on vital signs and five facet mindfulness questionnaire scores of college students. Journal of American College Health, 72(3), 905–913. 10.1080/07448481.2022.2060709

34. Doll, A., Hölzel, B. K., Mulej Bratec, S., Boucard, C. C., Xie, X., Wohlschläger, A. M., & Sorg, C. (2016). Mindful attention to breath regulates emotions via increased amygdala-prefrontal cortex connectivity. NeuroImage, 134, 305–313. 10.1016/j.neuroimage.2016.03.041

35. Fox, K. C. R., Dixon, M. L., Nijeboer, S., Girn, M., Floman, J. L., Lifshitz, M., Ellamil, M., Sedlmeier, P., & Christoff, K. (2016). Functional neuroanatomy of meditation: A review and meta-analysis of 78 functional neuroimaging investigations. Neuroscience and Biobehavioral Reviews, 65, 208–228. 10.1016/j.neubiorev.2016.03.021

36. Friston, K. J. (2010). The free-energy principle: A unified brain theory? Nature Reviews Neuroscience, 11(2), 127–138. 10.1038/nrn2787

37. Friston, K. J. (2019). Waves of prediction. PLoS Biology, 17(10), 1–7. 10.1371/journal.pbio.3000426

38. Galante, J., Friedrich, C., Dawson, A. F., Modrego-Alarcón, M., Gebbing, P., Delgado-Suárez, I., Gupta, R., Dean, L., Dalgleish, T., White, I. R., & Jones, P. B. (2021). Mindfulness-based programmes for mental health promotion in adults in nonclinical settings: A systematic review and meta-analysis of randomised controlled trials. In PLoS Medicine (Vol. 18, Issue 1). 10.1371/journal.pmed.1003481

39. Galante, J., Grabovac, A., Wright, M., Ingram, D. M., Van Dam, N. T., Sanguinetti, J. L., Sparby, T., van Lutterveld, R., & Sacchet, M. D. (2023). A Framework for the Empirical Investigation of Mindfulness Meditative Development. Mindfulness, 14(5), 1054–1067. 10.1007/s12671-023-02113-8

40. Gallant, S. N. (2016). Mindfulness meditation practice and executive functioning: Breaking down the benefit. Consciousness and Cognition, 40, 116–130. 10.1016/j.concog.2016.01.005

41. Ganesan, S., Beyer, E., Moffat, B., Van Dam, N. T., Lorenzetti, V., & Zalesky, A. (2022). Focused attention meditation in healthy adults: A systematic review and meta-analysis of cross-sectional functional MRI studies. Neuroscience and Biobehavioral Reviews, 141(May), 104846. 10.1016/j.neubiorev.2022.104846

42. Gao, J., Sun, R., Leung, H. K., Roberts, A., Wu, B. W. Y., Tsang, E. W., Tang, A. C. W., & Sik, H. H. (2023). Increased neurocardiological interplay after mindfulness meditation: a brain oscillation-based approach. Frontiers in Human Neuroscience, 17(June), 1–9. 10.3389/fnhum.2023.1008490

43. Gerritsen, R. J. S., & Band, G. P. H. (2018). Breath of Life: The Respiratory Vagal Stimulation Model of Contemplative Activity. Frontiers in Human Neuroscience, 12(February), 1–25. 10.3389/fnhum.2018.00397

44. Goheen, J., Anderson, J. A. E., Zhang, J., & Northoff, G. (2023). From Lung to Brain: Respiration Modulates Neural and Mental Activity. Neuroscience Bulletin. 10.1007/s12264-023-01070-5

45. Goheen, J., Wolman, A., Angeletti, L. L., Wolff, A., Anderson, J. A. E., & Northoff, G. (2024). Dynamic mechanisms that couple the brain and breathing to the external environment. Communications Biology, 7(1), 1–11. 10.1038/s42003-024-06642-3

46. Goldberg, S. B. (2022). A common factors perspective on mindfulness-based interventions. Nature Reviews Psychology, 1(10), 605–619. 10.1038/s44159-022-00090-8

47. Goldberg, S. B., Riordan, K. M., Sun, S., & Davidson, R. J. (2022). The Empirical Status of Mindfulness-Based Interventions: A Systematic Review of 44 Meta-Analyses of Randomized Controlled Trials. Perspectives on Psychological Science, 17(1), 108–130. 10.1177/1745691620968771

48. Guo, X., Wang, M., Wang, X., Guo, M., Xue, T., Wang, Z., Li, H., Xu, T., He, B., Cui, D., & Tong, S. (2022). Progressive increase of high-frequency EEG oscillations during meditation is associated with its trait effects on heart rate and proteomics: a study on the Tibetan Buddhist. *Cerebral Cortex (New York*, N.Y*. :* 1991), *32*(18), 3865–3877. 10.1093/cercor/bhab453

49. Haegens, S., Nácher, V., Luna, R., Romo, R., & Jensen, O. (2011). α-Oscillations in the monkey sensorimotor network influence discrimination performance by rhythmical inhibition of neuronal spiking. Proceedings of the National Academy of Sciences of the United States of America, 108(48), 19377–19382. 10.1073/pnas.1117190108

50. Harrison, O. K., Köchli, L., Marino, S., Luechinger, R., Hennel, F., Brand, K., Hess, A. J., Frässle, S., Iglesias, S., Vinckier, F., Petzschner, F. H., Harrison, S. J., & Stephan, K. E. (2021). Interoception of breathing and its relationship with anxiety. Neuron, 109(24), 4080–4093.e8. 10.1016/j.neuron.2021.09.045

51. Herrero, J. L., Khuvis, S., Yeagle, E., Cerf, M., & Mehta, A. D. (2018). Breathing above the brain stem: Volitional control and attentional modulation in humans. Journal of Neurophysiology, 119(1), 145–159. 10.1152/jn.00551.2017

52. Hsueh, B., Chen, R., Jo, Y. J., Tang, D., Raffiee, M., Kim, Y. S., Inoue, M., Randles, S., Ramakrishnan, C., Patel, S., Kim, D. K., Liu, T. X., Kim, S. H., Tan, L., Mortazavi, L., Cordero, A., Shi, J., Zhao, M., Ho, T. T., … Deisseroth, K. (2023). Cardiogenic control of affective behavioural state. Nature, 615(7951), 292–299. 10.1038/s41586-023-05748-8

53. Jelinčić, V., Van Diest, I., Torta, D. M., & von Leupoldt, A. (2022). The breathing brain: The potential of neural oscillations for the understanding of respiratory perception in health and disease. Psychophysiology, 59(5), 1–22. 10.1111/psyp.13844

54. Jensen, O., & Mazaheri, A. (2010). Shaping functional architecture by oscillatory alpha activity: Gating by inhibition. Frontiers in Human Neuroscience, 4(November), 1–8. 10.3389/fnhum.2010.00186

55. Jiang, H., He, B., Guo, X., Wang, X., Guo, M., Wang, Z., Xue, T., Li, H., Xu, T., Ye, S., Suma, D., Tong, S., & Cui, D. (2020). Brain-Heart Interactions Underlying Traditional Tibetan Buddhist Meditation. Cerebral Cortex (New York, N.Y. : 1991), 30(2), 439–450. 10.1093/cercor/bhz095

56. Kabat-Zinn, J. (2013). Full Catastrophe Living, revised edition: Using the Wisdom of Your Body and Mind to Face Stress, Pain, and Illness. Dell Publishing. 10.1037/032287

57. Kakumanu, R. J., Kumar, A., Venugopal, R., Sasidharan, A., Kumar, P., John, J. P., Mehrotra, S., Panth, R., & Kutty, B. M. (2018). Dissociating meditation pro fi ciency and experience dependent EEG changes during traditional Vipassana meditation practice. Biological Psychology, 135(September 2017), 65–75. 10.1016/j.biopsycho.2018.03.004

58. Kam, J. W. Y., Rahnuma, T., Park, Y. E., & Hart, C. M. (2022). Electrophysiological markers of mind wandering: A systematic review. NeuroImage, 258(June), 119372. 10.1016/j.neuroimage.2022.119372

59. Karunarathne, L. J. U., Amarasiri, W. A. D. L., & Fernando, A. D. A. (2024). Respiratory function in healthy long-term meditators: a systematic review. Systematic Reviews, 13(1), 1–14. 10.1186/s13643-023-02412-0

60. Kawashima, T., Shiratori, H., & Amano, K. (2024). The relationship between alpha power and heart rate variability commonly seen in various mental states. PLoS ONE, *19*(3 March), 1–14. 10.1371/journal.pone.0298961

61. Kerr, C. E., Sacchet, M. D., Lazar, S. W., Moore, C. I., & Jones, S. R. (2013). Mindfulness starts with the body: Somatosensory attention and top-down modulation of cortical alpha rhythms in mindfulness meditation. Frontiers in Human Neuroscience, 7(JAN), 1–15. 10.3389/fnhum.2013.00012

62. Kim, D. K., Lee, K. M., Kim, J., Whang, M. C., & Kang, S. W. (2013). Dynamic correlations between heart and brain rhythm during autogenic meditation. *Frontiers in Human Neuroscience*, JUL. 10.3389/fnhum.2013.00414

63. Klimesch, W. (2018). The frequency architecture of brain and brain body oscillations: an analysis. European Journal of Neuroscience, 48(7), 2431–2453. 10.1111/ejn.14192

64. Kluger, D. S., Balestrieri, E., Busch, N. A., & Gross, J. (2021). Respiration aligns perception with neural excitability. ELife, 10, 1–19. 10.7554/eLife.70907

65. Kluger, D. S., & Gross, J. (2021). Respiration modulates oscillatory neural network activity at rest. PLOS Biology, 19(11), e3001457. 10.1371/journal.pbio.3001457

66. Kluger, D. S., Gross, J., & Keitel, C. (2024). A Dynamic Link between Respiration and Arousal. Journal of Neuroscience, 44(47), 1–9. 10.1523/JNEUROSCI.1173-24.2024

67. Kriakous, S. A., Elliott, K. A., Lamers, C., & Owen, R. (2021). The Effectiveness of Mindfulness-Based Stress Reduction on the Psychological Functioning of Healthcare Professionals: a Systematic Review. Mindfulness, 12(1), 1–28. 10.1007/s12671-020-01500-9

68. Krishna, D., & Prasanna, K. (2023). Heart – brain Rhythmic Synchronization during Meditation : A Nonlinear Signal Analysis. 132–139. 10.4103/ijoy.ijoy

69. Laborde, S., Mosley, E., Thayer, J. F., & Quintana, D. S. (2017). Heart Rate Variability and Cardiac Vagal Tone in Psychophysiological Research – Recommendations for Experiment Planning , Data Analysis , and Data Reporting. Frontiers in Psychology, 8(February), 1–18. 10.3389/fpsyg.2017.00213

70. Lee, D. J., Kulubya, E., Goldin, P., Goodarzi, A., & Girgis, F. (2018). Review of the neural oscillations underlying meditation. Frontiers in Neuroscience, 12(MAR), 1–7. 10.3389/fnins.2018.00178

71. Lewis-Healey, E., Tagliazucchi, E., Canales-Johnson, A., & Bekinschtein, T. A. (2024). Breathwork-induced psychedelic experiences modulate neural dynamics. Cerebral Cortex, 34(8). 10.1093/cercor/bhae347

72. Li, X., Deng, J., Long, Y., Ma, Y., Wu, Y., Hu, Y., He, X., Yu, S., Li, D., Li, N., & He, F. (2024). Focus on brain-lung crosstalk: Preventing or treating the pathological vicious circle between the brain and the lung. Neurochemistry International, 178(January), 105768. 10.1016/j.neuint.2024.105768

73. Lieberman, J., McConnell, P., Estarellas, M., & Sacchet, M. (2024). Neurophysiological Mechanisms of Focused Attention Meditation: A Scoping Systematic Review. 10.31234/osf.io/rg7w6

74. Lomas, T., Ivtzan, I., & Fu, C. H. Y. (2015). A systematic review of the neurophysiology of mindfulness on EEG oscillations. Neuroscience and Biobehavioral Reviews, 57, 401–410. 10.1016/j.neubiorev.2015.09.018

75. Lovelace, J. W., Ma, J., & Augustine, V. (2024). Defining cardioception: Heart-brain crosstalk. Neuron, 1–4. 10.1016/j.neuron.2024.10.009

76. Lu, Y., & Rodriguez-larios, J. (2022). Nonlinear EEG signatures of mind wandering during breath focus meditation. Current Research in Neurobiology, 3(September), 100056. 10.1016/j.crneur.2022.100056

77. Lumma, A. L., Kok, B. E., & Singer, T. (2015). Is meditation always relaxing? Investigating heart rate, heart rate variability, experienced effort and likeability during training of three types of meditation. International Journal of Psychophysiology, 97(1), 38–45. 10.1016/j.ijpsycho.2015.04.017

78. Lutz, A., Jha, A. P., Dunne, J. D., & Saron, C. D. (2015). Investigating the phenomenological matrix of mindfulness-related practices from a neurocognitive perspective. American Psychologist, 70(7), 632–658. 10.1037/a0039585

79. Maric, V., Ramanathan, D., & Mishra, J. (2020). Respiratory regulation & interactions with neuro-cognitive circuitry. Neuroscience and Biobehavioral Reviews, 112(February 2019), 95–106. 10.1016/j.neubiorev.2020.02.001

80. Mazaheri, A., van Schouwenburg, M. R., Dimitrijevic, A., Denys, D., Cools, R., & Jensen, O. (2014). Region-specific modulations in oscillatory alpha activity serve to facilitate processing in the visual and auditory modalities. NeuroImage, 87, 356–362. 10.1016/j.neuroimage.2013.10.052

81. Mierau, A., Klimesch, W., & Lefebvre, J. (2017). State-dependent alpha peak frequency shifts: Experimental evidence, potential mechanisms and functional implications. Neuroscience, 360, 146–154. 10.1016/j.neuroscience.2017.07.037

82. Moeyersons, J., Amoni, M., Van Huffel, S., Willems, R., & Varon, C. (2019). R-DECO: an open-source Matlab based graphical user interface for the detection and correction of R-peaks. PeerJ Computer Science, 5(10), e226. 10.7717/peerj-cs.226

83. Moore, A., Gruber, T., Derose, J., & Malinowski, P. (2012). Regular, brief mindfulness meditation practice improves electrophysiological markers of attentional control. Frontiers in Human Neuroscience, 6(JANUARY 2012), 1–15. 10.3389/fnhum.2012.00018

84. Morales, J. F., Moeyersons, J., Armanac, P., Orini, M., Faes, L., Overeem, S., Van Gilst, M., Van Dijk, J., Huffel, S. Van, Bailon, R., & Varon, C. (2020). Model-Based Evaluation of Methods for Respiratory Sinus Arrhythmia Estimation. IEEE Transactions on Biomedical Engineering, 1–12. 10.1109/TBME.2020.3028204

85. Nakamura, N. H., Oku, Y., & Fukunaga, M. (2024). “Brain–breath” interactions: respiration-timing–dependent impact on functional brain networks and beyond. Reviews in the Neurosciences, 35(2), 165–182. 10.1515/revneuro-2023-0062

86. Oostenveld, R., Fries, P., Maris, E., & Schoffelen, J. M. (2011). FieldTrip: Open source software for advanced analysis of MEG, EEG, and invasive electrophysiological data. Computational Intelligence and Neuroscience, 2011. 10.1155/2011/156869

87. Palva, J. M., Palva, S., & Kaila, K. (2005). Phase synchrony among neuronal oscillations in the human cortex. Journal of Neuroscience, 25(15), 3962–3972. 10.1523/JNEUROSCI.4250-04.2005

88. Pernice, R., Antonacci, Y., Zanetti, M., Busacca, A., Marinazzo, D., Faes, L., & Nollo, G. (2021). Multivariate Correlation Measures Reveal Structure and Strength of Brain–Body Physiological Networks at Rest and During Mental Stress. Frontiers in Neuroscience, 14, 602584. 10.3389/fnins.2020.602584

89. Pion-Tonachini, L., Kreutz-Delgado, K., & Makeig, S. (2019). ICLabel: An automated electroencephalographic independent component classifier, dataset, and website. NeuroImage, 198, 181–197. 10.1016/j.neuroimage.2019.05.026

90. Pletzer, B., Kerschbaum, H., & Klimesch, W. (2010). When frequencies never synchronize: The golden mean and the resting EEG. Brain Research, 1335, 91–102. 10.1016/j.brainres.2010.03.074

91. Prätzlich, M., Kossowsky, J., Gaab, J., & Krummenacher, P. (2016). Impact of short-term meditation and expectation on executive brain functions. Behavioural Brain Research, 297, 268–276. 10.1016/j.bbr.2015.10.012

92. Prescott, S. L., & Liberles, S. D. (2022). Internal senses of the vagus nerve. Neuron, 110(4), 579–599. 10.1016/j.neuron.2021.12.020

93. Quach, D., Jastrowski Mano, K. E., & Alexander, K. (2016). A Randomized Controlled Trial Examining the Effect of Mindfulness Meditation on Working Memory Capacity in Adolescents. Journal of Adolescent Health, 58(5), 489–496. 10.1016/j.jadohealth.2015.09.024

94. Quintana, D. S., & Heathers, J. A. J. (2014). Considerations in the assessment of heart rate variability in biobehavioral research. Frontiers in Psychology, 5(JUL), 1–10. 10.3389/fpsyg.2014.00805

95. Rassi, E., Dorffner, G., Gruber, W., Schabus, M., & Klimesch, W. (2019). Coupling and Decoupling between Brain and Body Oscillations. Neuroscience Letters, 711(June), 134401. 10.1016/j.neulet.2019.134401

96. Richter, C. G., Babo-Rebelo, M., Schwartz, D., Tallon-Baudry, C., & Huizinga, J. D. (2017). Phase-amplitude coupling at the organism level: The amplitude of spontaneous alpha rhythm fluctuations varies with the phase of the infra-slow gastric basal rhythm. NeuroImage, 146, 951–958. 10.1016/j.neuroimage.2016.08.043

97. Ritz, T. (2024a). How to account for respiration in respiratory sinus arrhythmia: Publication standards for heart rate variability studies in Biological Psychology. Biological Psychology, 190. 10.1016/j.biopsycho.2024.108806

98. Ritz, T. (2024b). Putting back respiration into respiratory sinus arrhythmia or high-frequency heart rate variability: Implications for interpretation, respiratory rhythmicity, and health. Biological Psychology, 185(December 2023), 108728. 10.1016/j.biopsycho.2023.108728

99. Rodriguez-larios, J., & Alaerts, K. (2021). EEG alpha–theta dynamics during mind wandering in the context of breath focus meditation: An experience sampling approach with novice meditation practitioners. European Journal of Neuroscience, 53(6), 1855–1868. 10.1111/ejn.15073

100. Rodriguez-Larios, J., & Alaerts, K. (2019). Tracking Transient Changes in the Neural Frequency Architecture: Harmonic Relationships between Theta and Alpha Peaks Facilitate Cognitive Performance. The Journal of Neuroscience, 39(32), 6291–6298. 10.1523/jneurosci.2919-18.2019

101. Rodriguez-Larios, J., Bracho Montes de Oca, E. A., & Alaerts, K. (2021). The EEG spectral properties of meditation and mind wandering differ between experienced meditators and novices. NeuroImage, 245, 118669. 10.1016/j.neuroimage.2021.118669

102. Rodriguez-Larios, J., Faber, P., Achermann, P., Tei, S., & Alaerts, K. (2020). From thoughtless awareness to effortful cognition: alpha - theta cross-frequency dynamics in experienced meditators during meditation, rest and arithmetic. Scientific Reports, 10(1), 5419. 10.1038/s41598-020-62392-2

103. Rodriguez-Larios, J., Wong, K. F., & Lim, J. (2024). Assessing the effects of an 8-week mindfulness training program on neural oscillations and self-reports during meditation practice. PLoS ONE, *19*(6.0), 1–14. 10.1371/journal.pone.0299275

104. Rodriguez-Larios, J., Wong, K. F., Lim, J., & Alaerts, K. (2020). Mindfulness Training is Associated with Changes in Alpha-Theta Cross-Frequency Dynamics During Meditation. Mindfulness, 11(12), 2695–2704. 10.1007/s12671-020-01487-3

105. Rosenkranz, M. A., Lutz, A., Perlman, D. M., Bachhuber, D. R. W., Schuyler, B. S., MacCoon, D. G., & Davidson, R. J. (2016). Reduced stress and inflammatory responsiveness in experienced meditators compared to a matched healthy control group. Psychoneuroendocrinology, 68, 117–125. 10.1016/j.psyneuen.2016.02.013

106. Rusinova, A., Volodina, M., & Ossadtchi, A. (2024). Short-term meditation training alters brain activity and sympathetic responses at rest, but not during meditation. Scientific Reports, 14(1), 1–12. 10.1038/s41598-024-60932-8

107. Saggar, M., King, B. G., Zanesco, A. P., MacLean, K. A., Aichele, S. R., Jacobs, T. L., Bridwell, D. A., Shaver, P. R., Rosenberg, E. L., Sahdra, B. K., Ferrer, E., Tang, A. C., Mangun, G. R., Wallace, B. A., Miikkulainen, R., & Saron, C. D. (2012). Intensive training induces longitudinal changes in meditation state-related EEG oscillatory activity. Frontiers in Human Neuroscience, 6, 256. 10.3389/fnhum.2012.00256

108. Sargent, K. S., Martinez, E. L., Reed, A. C., Guha, A., Bartholomew, M. E., Diehl, C. K., Chang, C. S., Salama, S., Popov, T., Thayer, J. F., Miller, G. A., & Yee, C. M. (2024). Oscillatory Coupling Between Neural and Cardiac Rhythms. Psychological Science, 35(5), 517–528. 10.1177/09567976241235932

109. Sauseng, P., Klimesch, W., Gruber, W. R., & Birbaumer, N. (2008). Cross-frequency phase synchronization: A brain mechanism of memory matching and attention. NeuroImage, 40(1), 308–317. 10.1016/j.neuroimage.2007.11.032

110. Shaffer, F., & Ginsberg, J. P. (2017). An Overview of Heart Rate Variability Metrics and Norms. Frontiers in Public Health, 5, 258. 10.3389/fpubh.2017.00258

111. Short, E. B., Kose, S., Mu, Q., Borckardt, J., Newberg, A., George, M. S., & Kozel, F. A. (2010). Regional brain activation during meditation shows time and practice effects: An exploratory FMRI study. Evidence-Based Complementary and Alternative Medicine, 7(1), 121–127. 10.1093/ecam/nem163

112. Siebenhühner, F., Wang, S. H., Palva, J. M., & Palva, S. (2016). Cross-frequency synchronization connects networks of fast and slow oscillations during visual working memory maintenance. ELife, 5, 15–30. 10.7554/eLife.13451

113. Signorelli, C. M., Boils, J. D., Tagliazucchi, E., Jarraya, B., & Deco, G. (2022). From brain-body function to conscious interactions. Neuroscience and Biobehavioral Reviews, 141(August), 104833. 10.1016/j.neubiorev.2022.104833

114. Sik, H. H., Gao, J., Fan, J., Wai, B., Wu, Y., Leung, H. K., & Hung, Y. S. (2017). Using Wavelet Entropy to Demonstrate how Mindfulness Practice Increases Coordination between Irregular Cerebral and Cardiac Activities. *Journal of Visualized Experiments*, May, 1–10. 10.3791/55455

115. Silvani, A., Calandra-Buonaura, G., Dampney, R. A. L., & Cortelli, P. (2016). Brain–heart interactions: physiology and clinical implications. *Philosophical Transactions of the Royal Society A: Mathematical*, Physical and Engineering Sciences, 374(2067), 20150181. 10.1098/rsta.2015.0181

116. Soriano, J. R., Rodriguez-Larios, J., Varon, C., Castellanos, N., & Alaerts, K. (2024). Brain–Heart Interactions in Novice Meditation Practitioners During Breath Focus and an Arithmetic Task. Mindfulness. 10.1007/s12671-024-02431-5

117. Sparby, T., & Sacchet, M. D. (2022). Defining Meditation: Foundations for an Activity-Based Phenomenological Classification System. Frontiers in Psychology, 12(January). 10.3389/fpsyg.2021.795077

118. Sterling, P. (2018). Predictive regulation and human design. ELife, 7, 1–12. 10.7554/eLife.36133

119. Sugawara, A., Katsunuma, R., Terasawa, Y., & Sekiguchi, A. (2024). Interoceptive training impacts the neural circuit of the anterior insula cortex. Translational Psychiatry, 14(1), 1–7. 10.1038/s41398-024-02933-9

120. Sun, S., Hu, C., Pan, J., Liu, C., & Huang, M. (2019). Trait Mindfulness Is Associated With the Self-Similarity of Heart Rate Variability. Frontiers in Psychology, 10, 314. 10.3389/fpsyg.2019.00314

121. Tang, Y. Y., Ma, Y., Fan, Y., Feng, H., Wang, J., Feng, S., Lu, Q., Hu, B., Lin, Y., Li, J., Zhang, Y., Wang, Y., Zhou, L., & Fan, M. (2009). Central and autonomic nervous system interaction is altered by short-term meditation. Proceedings of the National Academy of Sciences of the United States of America, 106(22), 8865–8870. 10.1073/pnas.0904031106

122. Varon, C., Lazaro, J., Bolea, J., Hernando, A., Aguilo, J., Gil, E., Van Huffel, S., & Bailon, R. (2019). Unconstrained Estimation of HRV Indices after Removing Respiratory Influences from Heart Rate. IEEE Journal of Biomedical and Health Informatics, 23(6), 2386–2397. 10.1109/JBHI.2018.2884644

123. Walter, N., & Hinterberger, T. (2022). Determining states of consciousness in the electroencephalogram based on spectral , complexity , and criticality features. 8(1). 10.1093/nc/niac008

124. Wang, M. Y., Corcoran, A. W., McQueen, B., Freedman, G., Humble, G., Fitzgibbon, B. M., Fitzgerald, P. B., & Bailey, N. W. (2024). Experienced Meditators Show Enhanced Interaction Between Brain and Heart Functioning. Mindfulness, 15(12), 3198–3216. 10.1007/s12671-024-02482-8

125. Wielgosz, J., Schuyler, B. S., Lutz, A., & Davidson, R. J. (2016). Long-term mindfulness training is associated with reliable differences in resting respiration rate. Scientific Reports, 6(June), 1–6. 10.1038/srep27533

126. Wong, G. F., Sun, R., Adler, J., Yeung, K. W., Yu, S., & Gao, J. (2022). Loving-kindness meditation (LKM) modulates brain-heart connection: An EEG case study. Frontiers in Human Neuroscience, 16(September). 10.3389/fnhum.2022.891377

127. Xia, R., Chen, X., Engel, T. A., & Moore, T. (2024). Common and distinct neural mechanisms of attention. Trends in Cognitive Sciences, 28(6), 554–567. 10.1016/j.tics.2024.01.005

128. Yoshida, K., Takeda, K., Kasai, T., Makinae, S., Murakami, Y., Hasegawa, A., & Sakai, S. (2020). Focused attention meditation training modifies neural activity and attention : longitudinal EEG data in non-meditators. October 2019, 215–223. 10.1093/scan/nsaa020

129. Zaccaro, A., della Penna, F., Bubbico, F., Bayram, B., Parrotta, E., Perrucci, M. G., Costantini, M., & Ferri, F. (2025). Cardio-respiratory interactions in interoceptive perception: The role of heartbeat-modulated cortical oscillations. In *bioRxiv* (pp. 1–55). 10.1101/2025.04.14.648690

130. Zaccaro, A., Piarulli, A., Laurino, M., Garbella, E., Menicucci, D., Neri, B., & Gemignani, A. (2018). How Breath-Control Can Change Your Life: A Systematic Review on Psycho-Physiological Correlates of Slow Breathing. Frontiers in Human Neuroscience, 12(September), 1–16. 10.3389/fnhum.2018.00353

131. Zanetti, M., Faes, L., Nollo, G., De Cecco, M., Pernice, R., Maule, L., Pertile, M., & Fornaser, A. (2019). Information Dynamics of the Brain, Cardiovascular and Respiratory Network during Different Levels of Mental Stress. Entropy, 21(3), 275. 10.3390/e21030275

132. Zelano, C., Jiang, H., Zhou, G., Arora, N., Schuele, S., Rosenow, J., & Gottfried, J. A. (2016). Nasal respiration entrains human limbic oscillations and modulates cognitive function. Journal of Neuroscience, 36(49), 12448–12467. 10.1523/JNEUROSCI.2586-16.2016

133. Zsadanyi, S. E., Kurth, F., & Luders, E. (2021). The Effects of Mindfulness and Meditation on the Cingulate Cortex in the Healthy Human Brain: A Review. Mindfulness, 12(10), 2371–2387. 10.1007/s12671-021-01712-7

